# Matched marrow and blood profiling reveals compartment- and feature-specific liquid-biopsy signals in multiple myeloma

**DOI:** 10.64898/2026.09.09.26358882

**Authors:** Dor D. Abelman, Jenna Eagles, Stephanie Pedersen, David S. Scott, Arnavaz Danesh, Jeff P. Bruce, Stephenie D. Prokopec, Aimee Wong, Ellen N. Wei, Saumil Shah, Darrell White, Irwindeep Sandhu, Kevin Song, Alli Murugesan, Anthony Reiman, Suzanne Trudel, Trevor J. Pugh

## Abstract

Multiple myeloma is bone-marrow-predominant, motivating marrow plasma cell-free DNA (cfDNA) as a tumor-proximal liquid biopsy. We tested whether marrow plasma cfDNA better represents the myeloma genome than peripheral blood cfDNA by profiling matched marrow plasma cells and both cfDNA compartments from 74 patients across 372 cfDNA sample time points. Tumor fractions correlated between compartments (r=0.84), but marrow plasma cfDNA was not a superior surrogate: peripheral-blood cfDNA matched or exceeded marrow-plasma cfDNA for marrow-defined CNA and mutation recovery (77.4% vs 72.0%; 69.2% vs 65.4%), with high specificity. Disease-associated features were detected in 69/74 peripheral-blood and 68/74 marrow-plasma profiles. BCR clonotypes were the highest-recovery feature, with BM-dominant clonotypes recovered in 31/34 callable profiles per compartment. At high tumor fraction, adverse-lesion sensitivity reached 100% in both cfDNA compartments. Marrow plasma cfDNA showed distinct, handling-sensitive fragmentomic architecture (LOOCV AUC=0.910), demonstrating performance depends on compartment, biomarker class, tumor fraction and processing context.

## Introduction

Liquid-biopsy studies often ask whether cell-free DNA (cfDNA) recovers tumor alterations measured in a reference tissue. In multiple myeloma (MM), this comparison is not straightforward: the reference tissue is usually a single-site bone marrow aspirate, whereas cfDNA can be sampled from peripheral blood or bone marrow plasma. These compartments may differ in tumor fraction, anatomical representation, sample handling and background DNA composition. Concordance may also differ by analyte: copy-number alterations (CNAs), mutations, immunoglobulin (Ig) translocations, B-cell receptor (BCR) clonotypes and fragmentomic features have different biological interpretations, assay callability and detection thresholds. Thus, discordance between cfDNA and marrow plasma cells does not necessarily indicate liquid-biopsy failure; it may instead reflect the compartment sampled, the biomarker measured, the threshold applied, pre-analytical handling or uncertainty in the marrow reference itself.

MM provides a stringent setting for testing this problem. The disease is genomically heterogeneous, initiated in many cases by primary Ig translocations or hyperdiploidy and subsequently shaped by CNAs, somatic mutations, and clonal evolution.^1–3^ Yet diagnostic molecular profiling and post-treatment assessments remain primarily marrow-centred, including plasma cell phenotype and genomic evaluation, as well as measurable residual disease (MRD) assays, while imaging is used to assess focal or extramedullary disease.^4,5^ This dependence has practical and biological limits: myeloma involvement is spatially heterogeneous across the iliac crest, focal lesions, and extramedullary sites, and a single-site aspirate may under-represent regional subclones or total disease distribution.^6–8^ Marrow aspirates are also affected by hemodilution and sample-quality variation.^9,10^ cfDNA profiling offers a complementary route for molecular assessment of myeloma. Prior studies show that peripheral blood (PB) cfDNA can recover marrow-defined alterations, detect Ig rearrangements or structural events, reveal tumor alterations not captured in a sampled aspirate, and provide longitudinal readouts of response, progression, and clonal evolution.^11–17^ However, the same literature also shows that cfDNA and marrow are not interchangeable substrates. Sensitivity and concordance vary with tumor fraction, disease burden and distribution, lesion allele fraction, assay design, and treatment context.^14,15,18^ Blood-based clonotype and MRD assays are similarly context-dependent; they can be informative in selected settings, but they are not uniformly interchangeable with marrow MRD.^18–21^ These observations support the clinical promise of cfDNA as a complementary molecular source, while arguing against treating any single cfDNA source as a simple replacement for marrow profiling.

A matched cross-compartment benchmark is therefore needed to determine whether discordance reflects the cfDNA compartment, the analyte measured, tumor burden, assay calibration or uncertainty in the marrow reference. While BM plasma cells remain the clinical standard for genomic benchmarking, a single-site biopsy cannot fully capture the spatial complexity of myeloma biology.^4,6^ BM plasma cfDNA and PB cfDNA are related but non-identical molecular compartments, yet matched head-to-head data in myeloma remain limited. Their interchangeability should therefore be evaluated separately for mutations, CNAs, Ig translocations, BCR clonotypes, tumor fraction, and fragmentomic features. This feature-specific approach is especially important for fragmentomics. Fragment length, nucleosome-associated coverage patterns, genome-wide fragmentation profiles, and end motifs are biologically informative across cancers, but myeloma-specific evidence remains limited, and collection and processing conditions can influence fragmentomic readouts.^22–25^ The central question is therefore not whether PB cfDNA simply recapitulates marrow, but which molecular signals are concordant, complementary, or non-equivalent across compartments, and under what biological and technical conditions.

We therefore performed matched cross-compartment profiling of BM plasma cells, BM plasma cfDNA, and PB cfDNA across three myeloma cohorts. Using BM plasma cells as a clonal reference for genomic recovery, and applying prespecified feature-class-specific calibration, we evaluated which liquid-biopsy signals were concordant, complementary, or non-equivalent across compartments. This design separates the effects of sampling compartment, molecular feature class, tumor fraction, assay threshold, and reference definition, providing a framework for interpreting myeloma cfDNA as a calibrated molecular readout rather than a generic marrow surrogate.

## Results

### PB and BM plasma cfDNA show correlated tumor fractions but distinct molecular compositions

The paired-cfDNA cohort comprised 74 patients from three longitudinal studies: ALGONQUIN (n = 21), IMMAGINE (n = 11), and TFRIM4 (n = 42; Fig. 1A–B; Table 1). The cohort spanned newly diagnosed and relapsed disease settings with heterogeneous stage, subtype, and cytogenetic-risk distributions across studies (Table 1). Endpoint-specific analyses used the maximal evaluable subset for each assay, with denominators reported throughout.

**Figure 1.**
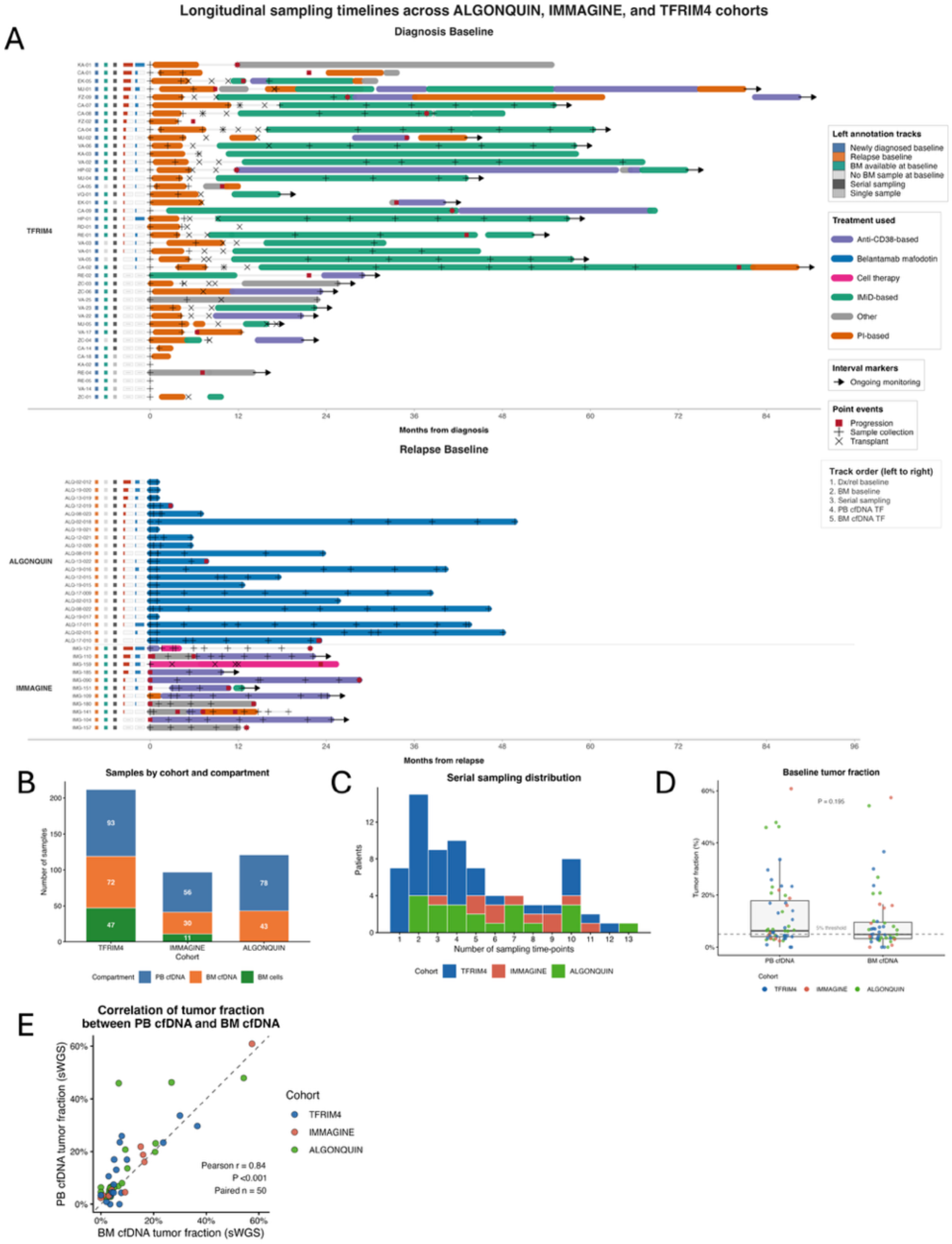
Longitudinal study design, sampling landscape, and baseline tumor-fraction concordance across BM plasma cfDNA and PB cfDNA compartments. **A**, Longitudinal sampling timelines across TFRIM4, ALGONQUIN, and IMMAGINE. Rows represent patients, intervals denote treatment classes, and symbols indicate sample collections, transplant, progression, and ongoing follow-up. Patients are grouped by baseline disease setting; time is anchored to diagnosis for newly diagnosed cases and to relapse for relapsed/refractory cases. Left annotation tracks summarize baseline disease setting, BM plasma-cell availability, serial sampling, and baseline PB cfDNA and BM plasma cfDNA tumor-fraction categories. **B**, Evaluable samples by cohort and compartment, showing the BM plasma-cell, BM plasma cfDNA, and PB cfDNA samples available for nested analyses. **C**, Number of distinct sampling time points per patient across the integrated cohort. **D**, Baseline tumor-fraction distributions in PB cfDNA and BM plasma cfDNA. Points represent samples, boxplots summarize distributions, and the dashed line marks the prespecified 5% tumor-fraction threshold used for stratified analyses. **E,** Paired baseline PB cfDNA and BM plasma cfDNA tumor fractions by shallow whole-genome sequencing (n = 51 pairs). The diagonal indicates equality, and on-panel labels report correlation statistics. Together, these panels define the cohort structure, analyte availability, longitudinal sampling density, and baseline tumor-fraction framework used for downstream analyses.

**Table 1.**
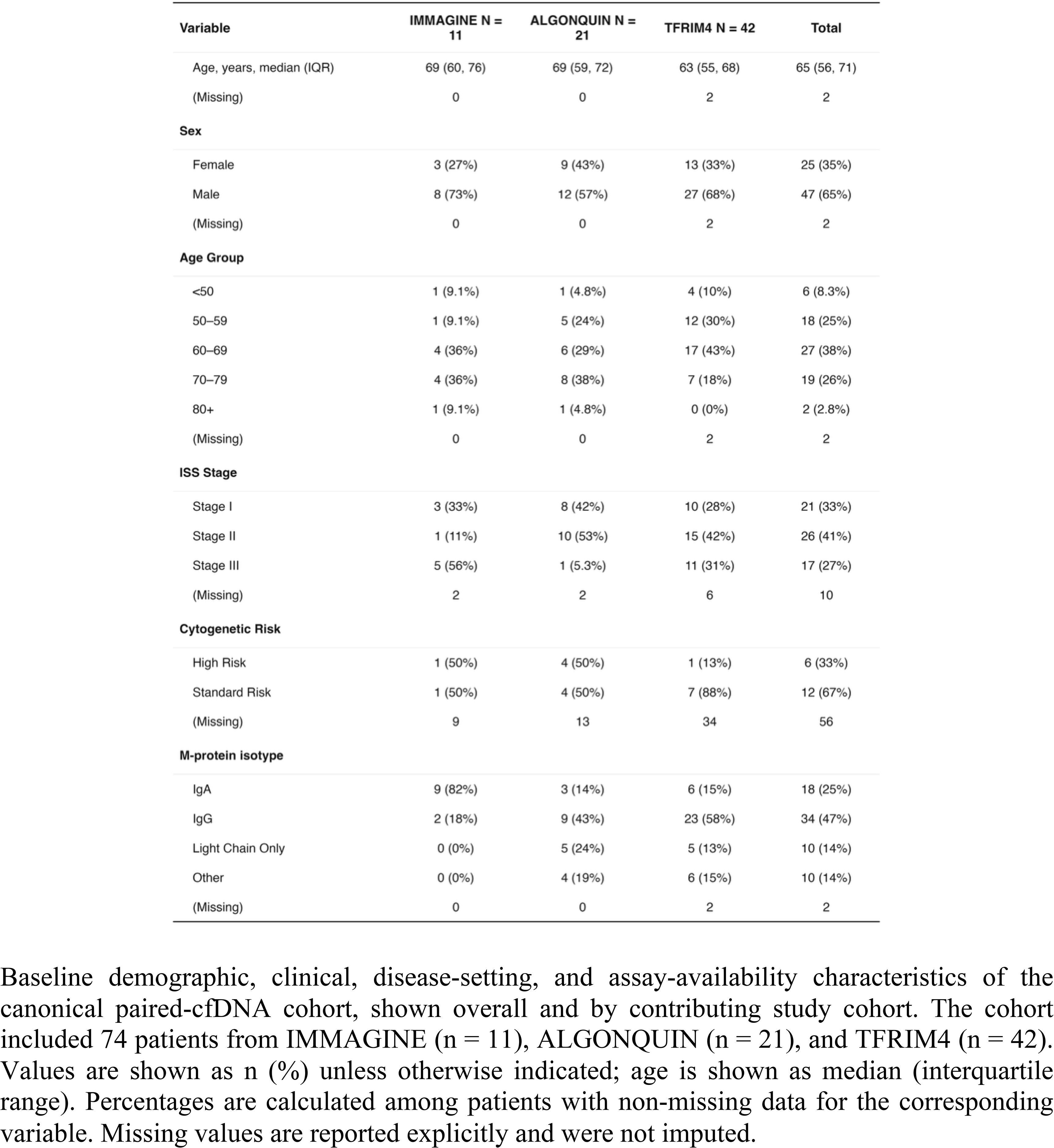
Baseline cohort characteristics and assay availability.

| Variable | IMMAGINE N =<br>11 | ALGONQUIN N =<br>21 | TFRIM4 N = 42 | Total |
| --- | --- | --- | --- | --- |
| Age, years, median (IQR) | 69 (60, 76) | 69 (59, 72) | 63 (55, 68) | 65 (56, 71) |
| (Missing) | 0 | 0 | 2 | 2 |
| **Sex** |  |  |  |  |
| Female | 3 (27%) | 9 (43%) | 13 (33%) | 25 (35%) |
| Male | 8 (73%) | 12 (57%) | 27 (68%) | 47 (65%) |
| (Missing) | 0 | 0 | 2 | 2 |
| **Age Group** |  |  |  |  |
| <50 | 1 (9.1%) | 1 (4.8%) | 4 (10%) | 6 (8.3%) |
| 50–59 | 1 (9.1%) | 5 (24%) | 12 (30%) | 18 (25%) |
| 60–69 | 4 (36%) | 6 (29%) | 17 (43%) | 27 (38%) |
| 70–79 | 4 (36%) | 8 (38%) | 7 (18%) | 19 (26%) |
| 80+ | 1 (9.1%) | 1 (4.8%) | 0 (0%) | 2 (2.8%) |
| (Missing) | 0 | 0 | 2 | 2 |
| **ISS Stage** |  |  |  |  |
| Stage I | 3 (33%) | 8 (42%) | 10 (28%) | 21 (33%) |
| Stage II | 1 (11%) | 10 (53%) | 15 (42%) | 26 (41%) |
| Stage III | 5 (56%) | 1 (5.3%) | 11 (31%) | 17 (27%) |
| (Missing) | 2 | 2 | 6 | 10 |
| **Cytogenetic Risk** |  |  |  |  |
| High Risk | 1 (50%) | 4 (50%) | 1 (13%) | 6 (33%) |
| Standard Risk | 1 (50%) | 4 (50%) | 7 (88%) | 12 (67%) |
| (Missing) | 9 | 13 | 34 | 56 |
| **M-protein isotype** |  |  |  |  |
| IgA | 9 (82%) | 3 (14%) | 6 (15%) | 18 (25%) |
| IgG | 2 (18%) | 9 (43%) | 23 (58%) | 34 (47%) |
| Light Chain Only | 0 (0%) | 5 (24%) | 5 (13%) | 10 (14%) |
| Other | 0 (0%) | 4 (19%) | 6 (15%) | 10 (14%) |
| (Missing) | 0 | 0 | 2 | 2 |

Baseline demographic, clinical, disease-setting, and assay-availability characteristics of the canonical paired-cfDNA cohort, shown overall and by contributing study cohort. The cohort included 74 patients from IMMAGINE (n = 11), ALGONQUIN (n = 21), and TFRIM4 (n = 42). Values are shown as n (%) unless otherwise indicated; age is shown as median (interquartile range). Percentages are calculated among patients with non-missing data for the corresponding variable. Missing values are reported explicitly and were not imputed.

Targeted-panel sequencing was available for PB cfDNA in 70 patients and BM plasma cfDNA in 60 patients (59 with both); 49 patients had matched BM tumor mutation data with matched-normal support, of which 36 had matched BM tumor translocation and immunoglobulin data. Baseline shallow whole-genome sequencing (sWGS) was available for PB cfDNA in 55 patients, BM plasma cfDNA in 51 patients, and BM plasma cells in 37 patients (51 patients with paired PB/BM plasma cfDNA sWGS, 18 with corresponding BM-cell sWGS; Extended Data Fig. 1A). Across follow-up, the 74 baseline patients contributed 372 cfDNA sample time points (227 PB cfDNA, 145 BM plasma cfDNA), with serial sampling in at least one compartment for 54/74 patients (Fig. 1B–C). Attrition reflected insufficient CD138+ BM plasma cells, absence of matched-normal support, and library or sequencing failures. Thirteen TFRIM4 baseline BM plasma samples (13/69) failed library QC. These samples were characterized by prominent adapter peaks and an absence of detectable cfDNA insert peaks following library preparation. This pattern is consistent with poor recoverable cfDNA quality after overnight EDTA shipment without cell-stabilizing preservatives. In contrast, locally processed IMMAGINE BM plasma cfDNA succeeded in all 38/38 evaluable cases (Extended Data Fig. 1A,D).

PB and BM plasma cfDNA shared a tumor-burden axis but were not interchangeable. Among 51 paired baseline tumor-fraction estimates, tumor fractions were strongly correlated (Pearson r = 0.84, P = 1.07 × 10^-14; Spearman ρ = 0.77, P = 2.92 × 10^-11; Fig. 1E). PB cfDNA more often exceeded the 5% high-tumor-fraction threshold than BM plasma cfDNA (33/55 PB versus 25/51 BM plasma cfDNA samples; Fig. 1D). This shared axis was modified by fragment-size composition and sample quality. In the BM plasma cfDNA compartment, the proportion of long fragments (221-400 bp) was negatively correlated with ichorCNA tumor fraction (Pearson r = - 0.24, P = 0.009; Extended Data Fig. 1E), consistent with dilution by non-tumor genomic DNA or ex vivo leukocyte lysis in shipped marrow samples rather than a purely biological difference in tumor shedding. Median baseline cfDNA yields were 356.6 ng in PB and 279 ng in BM plasma (Extended Data Fig. 1B; Supplementary Table 1).

### BM plasma cfDNA has distinct fragmentation and end-motif profiles

We first asked whether PB cfDNA and BM plasma cfDNA behaved as interchangeable liquid-biopsy analytes or retained compartment-specific molecular features. In matched baseline cfDNA pairs (n = 51), BM plasma cfDNA showed broader fragment-size remodeling than PB cfDNA, with attenuation of the canonical mono-nucleosomal peak and redistribution into both shorter and longer fragment-size ranges rather than a simple short-fragment enrichment (Fig. 2A; Extended Data Fig. 2A). Compared with PB cfDNA, BM plasma cfDNA had a higher short-fragment fraction (<150 bp; 24.4% vs 17.9%, paired Wilcoxon P < 0.001), slightly larger median insert size (170 vs 168 bp, P = 0.0046), greater enrichment of 100–149 bp fragments (20.2% vs 17.2%, P = 0.0014), and markedly greater enrichment of 221–400 bp fragments (16.4% vs 9.3%, P = 1.7 × 10⁻⁸) (Fig. 2B,C; Supplementary Table 2). BM plasma cfDNA also showed lower 150–180 bp mono-nucleosomal representation, lower mono:di-nucleosome ratio, and higher insert-size entropy (Extended Data Fig. 2A).

**Figure 2.**
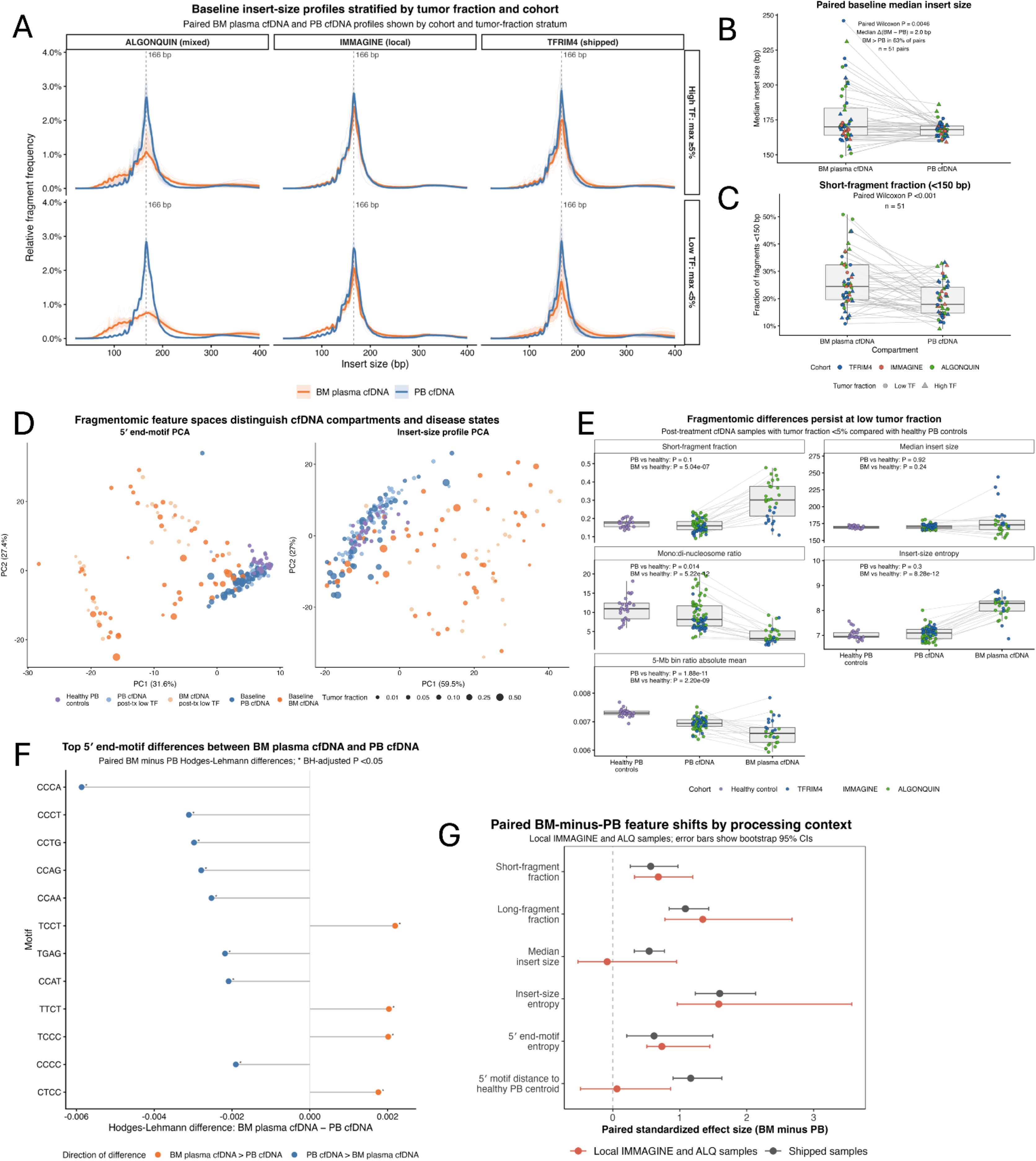
BM plasma cfDNA and PB cfDNA have distinct fragmentomic profiles that are sensitive to processing context. **A**, Baseline insert-size profiles for paired BM plasma cfDNA and PB cfDNA samples, stratified by cohort and pair-level tumor-fraction category. Individual profiles and compartment medians are shown; dashed lines mark the mono-nucleosomal peak. **B,** Paired baseline median insert size in BM plasma cfDNA and PB cfDNA, with lines connecting matched patient samples. **C,** Paired baseline short-fragment fraction (<150 bp) in matched BM plasma cfDNA and PB cfDNA samples. **D,** Principal-component analyses of 5′ end-motif profiles and insert-size profiles. Points represent healthy PB controls, low-tumor-fraction reference samples, and baseline PB cfDNA and BM plasma cfDNA samples; point size denotes tumor fraction for patient-derived samples. **E,** Fragmentomic features in healthy PB controls and low-tumor-fraction post-baseline PB cfDNA and BM plasma cfDNA samples, with matched cfDNA pairs connected where available. **F,** Top 5′ end motifs ranked by paired BM plasma cfDNA minus PB cfDNA differences, showing the direction and magnitude of compartment-specific motif shifts. On-panel labels report paired tests, healthy-control comparisons, or false-discovery-adjusted motif results, as appropriate. **G,** Processing-context sensitivity analysis of paired BM plasma cfDNA minus PB cfDNA fragmentomic feature shifts. Points show paired effect sizes and error bars show patient-bootstrap 95% confidence intervals, stratified by BM plasma processing context. Comparisons of paired deltas did not remain significant after Benjamini-Hochberg adjustment. Together, these analyses show that BM plasma cfDNA and PB cfDNA differ in fragment-size architecture and 5′ end-motif composition, including in low-tumor-fraction settings, supporting compartment-specific interpretation rather than a single tumor-fraction-scaled cfDNA model.

Several observations argued against a purely tumor-fraction-driven explanation for the PB versus BM plasma cfDNA fragmentomic differences. First, in tumor-fraction-stratified contrasts, BM plasma cfDNA showed broad redistribution around the mono-nucleosomal peak, whereas PB cfDNA retained a more canonical mono-nucleosomal profile (Extended Data Fig. 2C). Second, post-treatment low-tumor-fraction PB cfDNA closely resembled healthy PB cfDNA controls across primary fragment-size features, whereas low-tumor-fraction BM plasma cfDNA remained shifted, with higher short-fragment fraction, higher insert-size entropy, lower mono:di-nucleosome ratio, and lower 5-Mb fragmentation-ratio absolute mean (Fig. 2D,E). PCA similarly separated low-tumor-fraction BM plasma cfDNA from low-tumor-fraction PB cfDNA and healthy PB controls (Fig. 2D; Extended Data Fig. 2D). Third, 5′ end-motif analyses showed concordant compartment-associated shifts, including depletion of CCCA, CCTG, CCAG, CCCT, and related C-rich motifs and enrichment of selected T-rich motifs in BM plasma cfDNA relative to PB cfDNA, with dominant paired effects significant after false-discovery correction (Fig. 2F). Related end-motif summary features remained shifted in low-tumor-fraction BM plasma cfDNA relative to healthy PB controls and low-tumor-fraction PB cfDNA, and high-versus-low tumor-fraction motif contrasts showed compartment-specific patterns (Extended Data Fig. 3A,B).

Finally, a leave-one-patient-out logistic-regression classifier using fragment-size and 5′ end-motif summary features distinguished BM plasma cfDNA from PB cfDNA across matched baseline pairs (LOOCV AUC, 0.910; 95% CI, 0.842–0.978; within-patient label-permutation P = 0.001; Extended Data Fig. 3C). Performance was retained in low-and high-tumor-fraction subsets (AUC, 0.856 and 0.872, respectively) and study-stratified analyses showed consistent compartment separation across contributing cohorts, with the weakest separation in IMMAGINE (Extended Data Fig. 3C-F; Supplementary Notes). Together, these results support quantifiable compartment separability across tumor-fraction strata and study contexts.

We next asked whether the BM plasma cfDNA fragmentomic profile was explained primarily by pre-separation shipment. In paired baseline BM-minus-PB contrasts, local/non-shipped BM plasma cfDNA appeared closer to matched PB cfDNA than overnight-shipped BM plasma cfDNA for several handling-sensitive features, particularly long-fragment fraction and 5′ end-motif distance to the healthy-PB centroid (Fig. 2A,G; Extended Data Fig. 1F; Extended Data Fig. 2C). However, local/non-shipped BM plasma cfDNA did not collapse onto PB cfDNA. In strict local/non-shipped matched baseline pairs (n = 11), BM plasma cfDNA retained significant shifts in short-fragment fraction, 221–400 bp long-fragment fraction, 150–180 bp mono-nucleosomal representation, mono:di-nucleosome ratio, insert-size entropy and 5′ end-motif entropy (paired Wilcoxon P = 0.023, 0.007, 0.007, 0.005, 0.004 and 0.011, respectively; Supplementary Table 2, Extended Data Fig. 2B). In overnight-shipped matched pairs (n = 40), the same core feature directions were observed, with larger point estimates for several features, including short-fragment fraction, 221–400 bp fraction, loss of 150–180 bp mono-nucleosomal representation, insert-size entropy and 5′ end-motif entropy (all paired P ≤ 0.001; Fig. 2G; Supplementary Table 2). Local-versus-shipped comparisons of paired deltas did not remain significant after Benjamini–Hochberg correction. Overall, BM plasma cfDNA and PB cfDNA shared some fragmentomic structure but showed systematic differences in fragment size and end-motif composition, supporting compartment- and processing-aware interpretation rather than normalization to a single healthy peripheral-blood cfDNA reference.

### Genomic recovery from cfDNA is feature-class-specific

We benchmarked baseline cfDNA genomic recovery against matched BM CD138+ tumor cells. Because mutations, CNAs, translocations, and BCR clonotypes differ in signal structure and assay callability, each biomarker class was evaluated under a calibrated primary framework (Methods; Supplementary Notes). Tier 1/Tier 2 mutation thresholds and BM-reference stability were calibrated using endpoint-specific performance, driver sensitivity and cfDNA-only burden criteria (Extended Data Fig. 4A–F; Supplementary Tables 3 and 4). CNA, translocation and BCR frameworks were calibrated separately: CNA framework selection and callability were evaluated across candidate arm-level rules (Extended Data Fig. 6A,B; Supplementary Table 10), translocation thresholds and rescue rules were benchmarked against BM plasma-cell and FISH-referenced endpoints (Extended Data Fig. 6C–E; Supplementary Table 11), and BCR target-locus callability, BM-matched clonotype support and recovery thresholds were assessed across target-read and clonotype-read cutoffs (Extended Data Fig. 6F–H; Supplementary Table 12).

Under these calibrated definitions, the cross-compartment oncoprint showed shared disease architecture across BM tumor cells, PB cfDNA, and BM plasma cfDNA, together with compartment-specific gains and losses (Fig. 3A). CNA recovery was the strongest lesion-specific cfDNA readout: relative to BM tumor cells, PB cfDNA recovered BM-reference CNA events with 77.4% sensitivity (24/31 events) and 98.7% specificity (across 110 callable event tests), while BM plasma cfDNA recovered CNAs with 72.0% sensitivity (18/25) and 92.3% specificity (across 90 callable tests; Fig. 3B–C; Table 2). Mutation recovery was less sensitive but highly specific in both compartments (PB cfDNA 69.2% sensitivity / 99.4% specificity; BM plasma cfDNA 65.4% / 98.9%). Apparent cfDNA performance therefore depended strongly on biomarker class, with CNAs recovered more consistently than point mutations under the primary framework.

**Figure 3.**
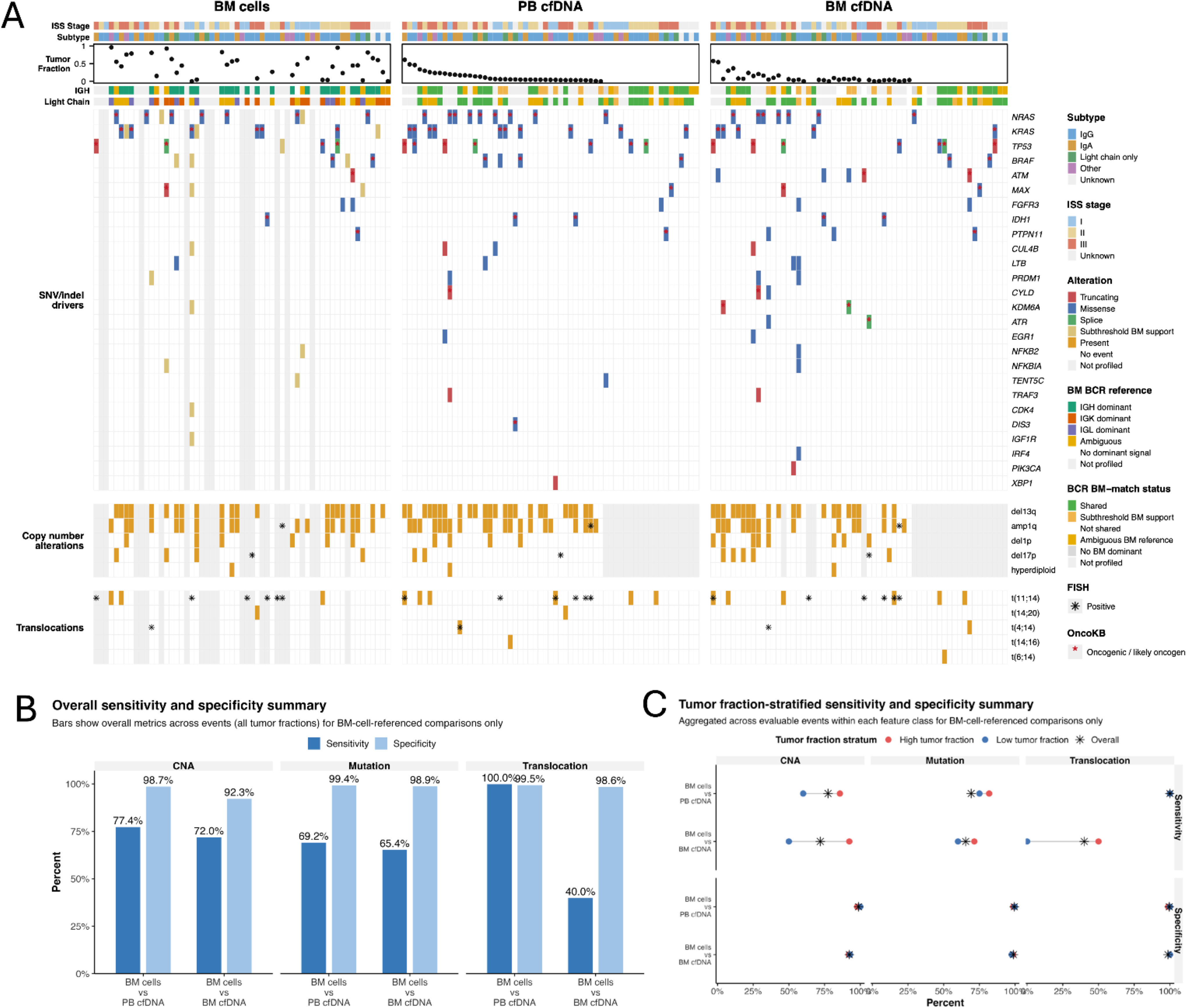
PB cfDNA and BM plasma cfDNA recover BM plasma-cell-defined genomic features with endpoint-specific performance. **A**, Baseline three-compartment oncoprint aligned by patient across BM plasma cells, PB cfDNA, and BM plasma cfDNA. The displayed targeted-panel cohort includes 59 paired PB cfDNA and BM plasma cfDNA cases, of which 47 also had matched baseline BM plasma-cell profiling. Rows show recurrent driver mutations, canonical copy-number alterations, key translocations, and BCR features, allowing shared and compartment-specific genomic architecture to be inspected within each patient. **B,** BM plasma-cell-referenced sensitivity and specificity for copy-number alterations, mutations, and translocations in PB cfDNA and BM plasma cfDNA. Bars summarize aggregate event-level performance under the calibrated primary frameworks. Sensitivity denominators are BM-cell-positive events and specificity denominators are BM-cell-negative events. For BM cells versus PB cfDNA, sensitivity/specificity denominators were 31/79 for CNAs, 26/2039 for mutations, and 5/217 for translocations; for BM cells versus BM plasma cfDNA, denominators were 25/65, 26/2039, and 5/217, respectively. Translocation sensitivity is based on five BM plasma-cell-reference-positive events and should be interpreted in the context of the small event set. **C,** Tumor-fraction-stratified BM plasma-cell-referenced sensitivity and specificity by feature class and cfDNA compartment. Points summarize event-level recovery within each calibrated endpoint; high- and low-tumor-fraction sensitivity/specificity denominators were, respectively, 21/44 and 10/35 for PB CNA, 11/619 and 4/416 for PB mutation, 2/76 and 2/46 for PB translocation, 13/37 and 12/28 for BM plasma CNA, 7/518 and 5/415 for BM plasma mutation, and 2/58 and 1/41 for BM plasma translocation. Together, these analyses show that both PB cfDNA and BM plasma cfDNA recover marrow-defined genomic features, but performance depends on biomarker class, tumor fraction, and calibration framework.

**Table 2.**
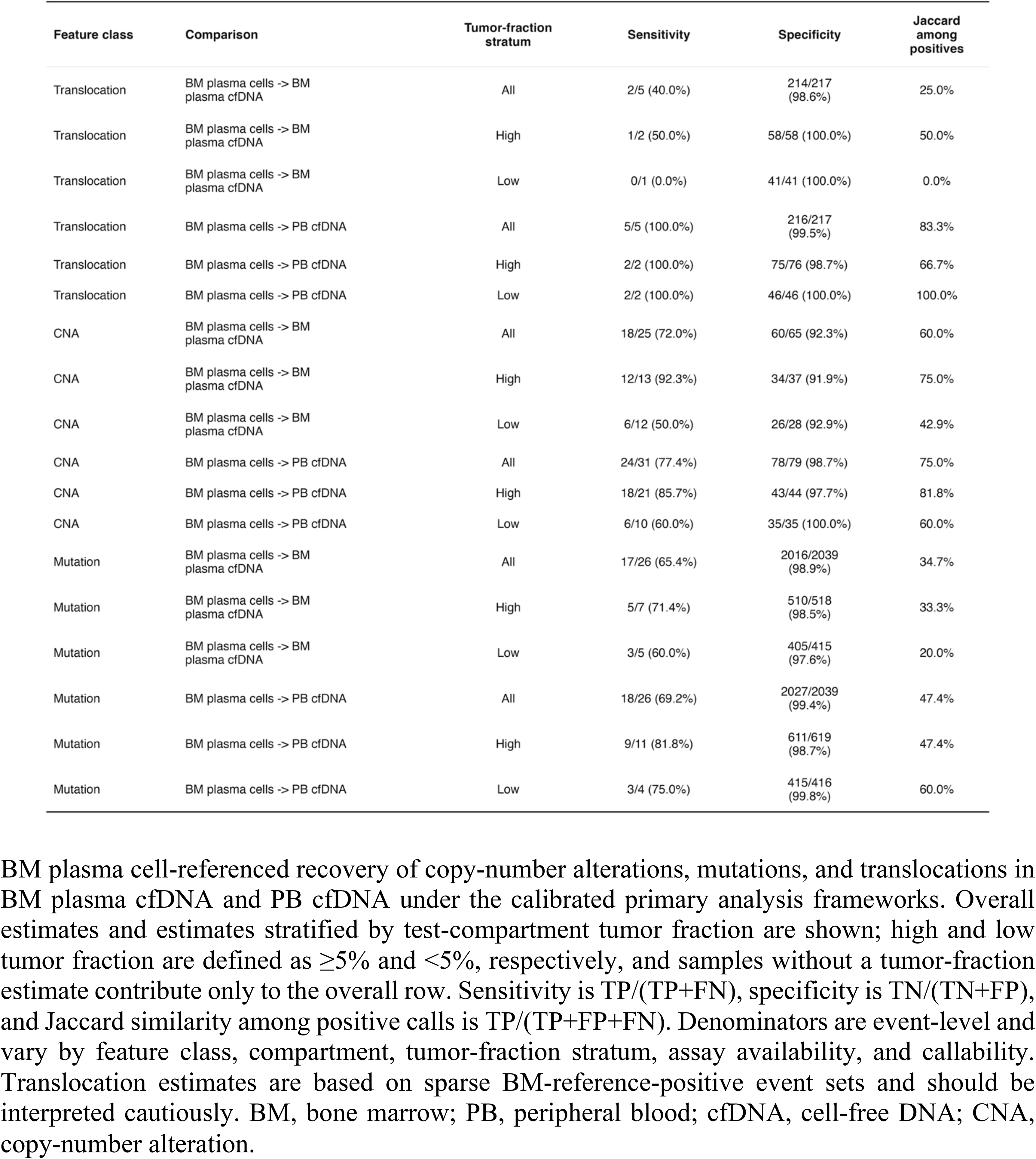
BM plasma cell-referenced molecular recovery by compartment.

Translocation recovery was informative but event-limited. Among five BM-reference–positive translocation events, PB cfDNA recovered all five and BM plasma cfDNA recovered two, with a specificity of 99.5% (PB) and 98.6% (BM plasma) across 222 callable event tests. However, BM-reference–positive translocation events were event-limited and should not be interpreted as a stable compartment hierarchy. Orthogonal FISH-referenced event-level benchmarking was similarly denominator-limited (Extended Data Fig. 6E; Supplementary Table 11). BCR recovery represented a distinct clonotype-based feature class, evaluated as sample-level detectability within the BM-dominant callable subset (Fig. 4A–B). cfDNA recovery is feature-class-specific, requiring separate calibration for each readout rather than a single aggregate concordance metric.

**Figure 4.**
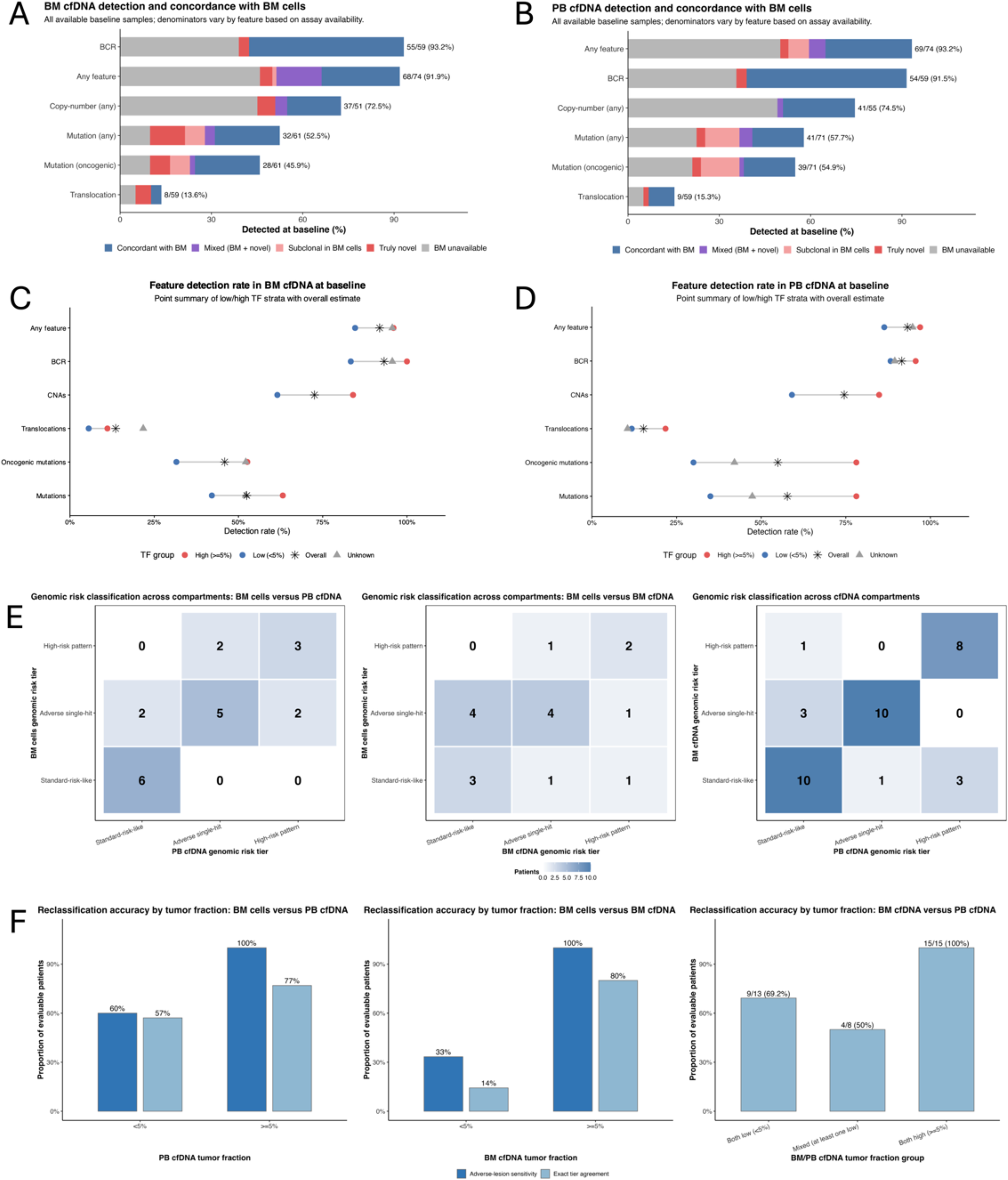
Feature detection sensitivity and genomic-risk concordance across BM plasma cfDNA and PB cfDNA compartments. **A**, Baseline BM plasma cfDNA feature detection across evaluable endpoint classes. Bars show the proportion of evaluable samples with BCR, any genomic feature, any copy-number alteration, any mutation, any oncogenic mutation, or translocation detected in BM plasma cfDNA. Stacked segments indicate whether detected features were concordant with BM plasma cells, mixed BM-supported plus novel, subclonal in BM plasma cells, novel, or lacked an available BM plasma-cell reference. Labels report detected/evaluable counts and percentages. **B,** Baseline PB cfDNA feature detection and BM plasma-cell concordance, displayed as in A. **C,** Baseline BM plasma cfDNA feature-detection rates among 74 baseline patients, stratified by BM plasma cfDNA tumor fraction, with overall rates shown for comparison: high ≥5% (n = 25), low <5% (n = 26), and unknown (n = 23). **D,** Baseline PB cfDNA feature-detection rates among the same 74 patients, stratified by PB cfDNA tumor fraction: high ≥5% (n = 33), low <5% (n = 22), and unknown (n = 19). Feature-specific denominators vary by assay availability and callability: any feature, n = 74; mutations/oncogenic mutations, n = 61 BM and n = 71 PB; translocations, n = 59; CNAs, n = 51 BM and n = 55 PB; and BCR, n = 59. **E,** Pairwise genomic-risk classification heat maps comparing BM plasma cells with PB cfDNA, BM plasma cells with BM plasma cfDNA, and BM plasma cfDNA with PB cfDNA. Cells report the number of patients assigned to each combination of three-level genomic-risk tiers: standard-risk-like, adverse single-hit, or high-risk pattern. **F,** Genomic-risk reclassification accuracy by tumor fraction. Together, these analyses show that baseline cfDNA recovery is feature-class and compartment dependent, with high BCR and broad-feature detection, lower translocation recovery, and more reliable genomic-risk classification when tumor fraction is sufficient in the queried cfDNA compartment.

### cfDNA-only mutations include subthreshold marrow signal

Mutation discordance required threshold-aware adjudication rather than binary classification. After calibration, recurrent low-confidence cfDNA-only patterns were demoted from Tier 1 to an exploratory tier (Extended Data Fig. 5A–B; Supplementary Tables 5–6). Among residual cfDNA-only Tier 1 calls, 9/12 PB cfDNA–only mutations and 5/23 BM plasma cfDNA–only mutations showed exact subthreshold support in matched BM tumor cells below the 10% reference cutoff (Extended Data Fig. 5C–E; Supplementary Table 7). In a broader lookback across 72 discordant cfDNA mutation observations (which include cfDNA-only Tier 1 calls plus other discordant categories adjudicated in Supplementary Table 8), 15/72 (20.8%) showed additional subthreshold BM evidence (Supplementary Tables 8–9). SBS-class summaries did not show significant Tier 1 compartment-specific spectrum differences (Extended Data Fig. 5F–G). Residual cfDNA-only calls were therefore adjudicated as possible low-VAF marrow signal, spatial or compartmental heterogeneity, or technical noise rather than automatically classified as false positives (Supplementary Notes).

### Detection scales with tumor fraction in a class-specific manner

Baseline cfDNA profiling detected disease-associated features in most patients, but detectability varied across biomarker classes, compartments, and tumor fractions. At least one assay-defined baseline cfDNA feature was detected in 69/74 PB cfDNA profiles (93.2%) and 68/74 BM plasma cfDNA profiles (91.9%). In the broad baseline feature-detection inventory, PB cfDNA detected a BCR signal in 54/59 profiles (91.5%), CNAs in 41/55 (74.5%), any mutation in 41/71 (57.7%), oncogenic mutation in 39/71 (54.9%), and translocations in 9/59 (15.3%) (Fig. 4A,B). BM plasma cfDNA detected a BCR signal in 55/59 profiles (93.2%), CNAs in 37/51 (72.5%), any mutation in 32/61 (52.5%), oncogenic mutation in 28/61 (45.9%), and translocations in 8/59 (13.6%) (Fig. 4A,B). In the stricter BM-dominant callable clonotype endpoint, the matched BM-dominant clonotype was recovered in 31/34 PB cfDNA and 31/34 BM plasma cfDNA profiles (Extended Data Fig. 6F–H; Supplementary Table 12).

Tumor fraction shaped lesion-specific detection differently across cfDNA compartments. In BM plasma cfDNA, any-mutation detection increased from 8/19 (42.1%) below the 5% tumor-fraction threshold to 12/19 (63.2%) at or above 5%, while oncogenic-mutation detection increased from 6/19 (31.6%) to 10/19 (52.6%). CNA detection increased from 16/26 (61.5%) to 21/25 (84.0%), and BCR recovery from 15/18 (83.3%) to 18/18 (100.0%) (Fig. 4C). In PB cfDNA, the tumor-fraction gradient was more pronounced: any-mutation detection increased from 7/20 (35.0%) to 25/32 (78.1%), oncogenic-mutation detection from 6/20 (30.0%) to 25/32 (78.1%), CNA detection from 13/22 (59.1%) to 28/33 (84.8%), and any-feature detection from 19/22 (86.4%) to 32/33 (97.0%). BCR recovery remained high across PB tumor-fraction strata, increasing from 15/17 (88.2%) to 22/23 (95.7%) (Fig. 4D). Thus, cfDNA detectability was feature- and compartment-dependent rather than reducible to a single sensitivity estimate.

Genomic-risk classification provided a more clinically interpretable endpoint within a smaller evaluable subset. Exact three-tier agreement was 70.0% (14/20) for BM tumor cells versus PB cfDNA, 52.9% (9/17) for BM tumor cells versus BM plasma cfDNA, and 77.8% (28/36) for BM plasma cfDNA versus PB cfDNA, with higher recovery of binary adverse-lesion status than exact tier labels in BM-reference comparisons (Fig. 4E). Reclassification improved with higher tumor fraction, particularly in BM-reference comparisons: exact tier agreement rose from 57% to 77% for PB cfDNA and from 14% to 80% for BM plasma cfDNA, and adverse-lesion sensitivity from 60% to 100% and 33% to 100%, respectively (Fig. 4F). These data support baseline cfDNA as a broadly informative molecular readout, while defining the tumor-fraction-dependent limits of lesion-specific detection and genomic-risk classification.

### Baseline cfDNA burden shows exploratory clinical associations in newly diagnosed myeloma

We next assessed whether baseline cfDNA burden related to progression-free survival (PFS) in TFRIM4, a uniformly newly diagnosed cohort with mature follow-up. In univariable Cox models, continuous baseline cfDNA feature burden was associated with shorter PFS for both PB cfDNA (n = 39, 17 events; HR 1.91 per feature, 95% CI 1.11–3.29; P = 0.020) and BM plasma cfDNA (HR 2.38 per feature, 95% CI 1.22–4.63; P = 0.011; Fig. 5C). Dichotomized exploratory Kaplan– Meier comparisons (0–1 vs ≥2 detected feature classes) were directionally consistent but underpowered (PB: HR 2.13, 95% CI 0.69–6.58, log-rank P = 0.179; BM plasma: HR 3.15, 95% CI 0.90–11.04, P = 0.060; Fig. 5A–B). In the complete-case subset with ISS, PB cfDNA burden, and BM plasma cfDNA burden available (n = 36, 17 events), adding cfDNA burden to ISS-only models did not reach significance (LR-test P = 0.117 for PB; P = 0.061 for BM plasma; Fig. 5D). Baseline cfDNA feature burden also increased with advancing International Staging System (ISS) stage in both compartments (Kruskal–Wallis P = 0.016 PB; P = 0.019 BM plasma; Fig. 5E). These exploratory associations support clinical coherence and motivate prospective validation but do not establish independent prognostic value in this cohort.

**Figure 5.**
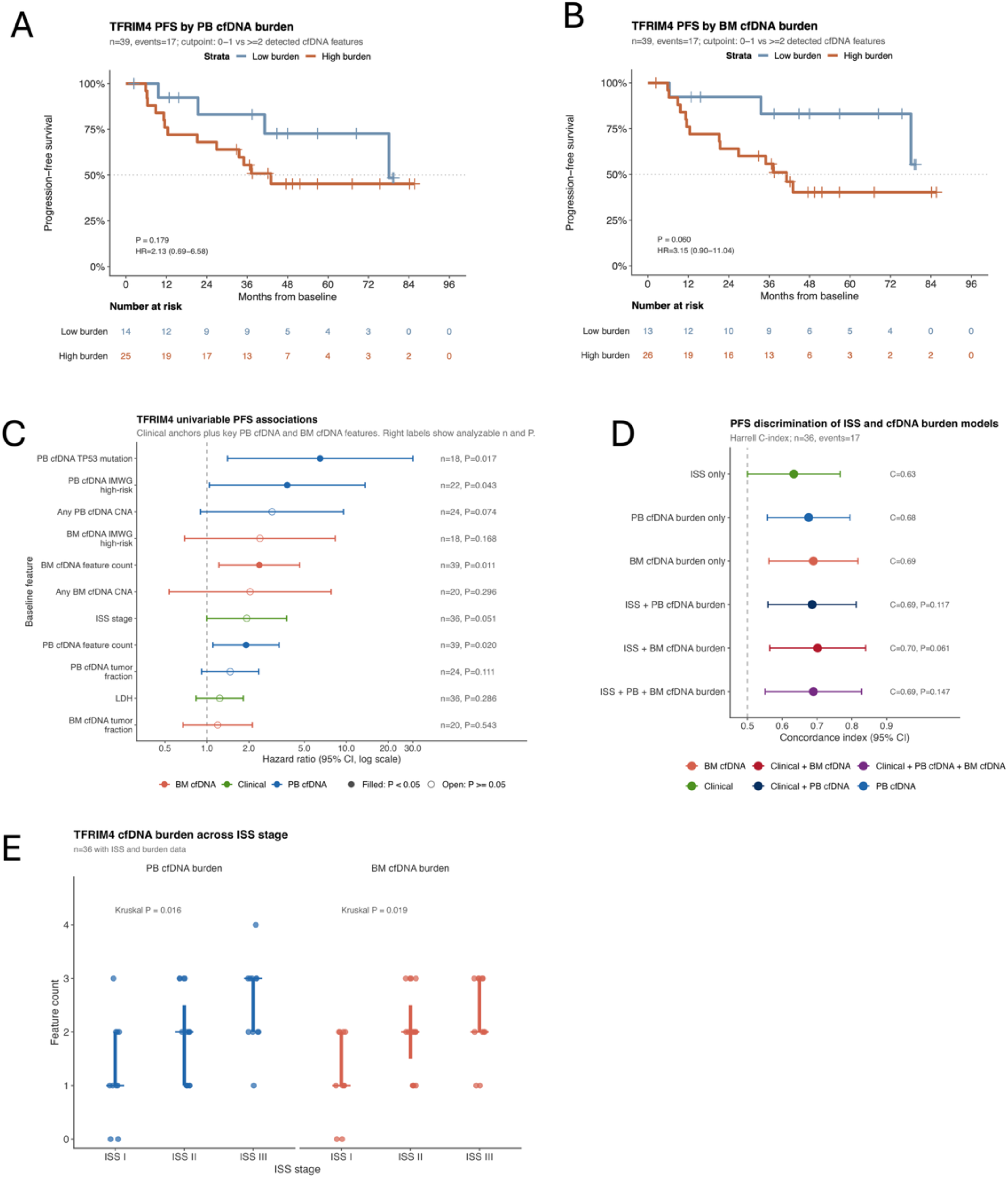
Baseline cfDNA burden shows exploratory associations with progression-free survival and ISS stage in TFRIM4. **A**, Kaplan-Meier estimates of progression-free survival in TFRIM4 stratified by baseline PB cfDNA burden, defined as 0-1 versus ≥2 detected cfDNA feature classes. The analysis included 39 patients and 17 PFS events; the low-burden group included 14 patients with 4 events, and the high-burden group included 25 patients with 13 events. The hazard ratio is from a Cox model comparing high versus low burden, and the P value is from a log-rank test. **B,** Kaplan-Meier estimates of progression-free survival stratified by baseline BM plasma cfDNA burden using the same burden definition. The low-burden group included 13 patients with 3 events, and the high-burden group included 26 patients with 14 events. **C, F**orest plot of univariable Cox associations between selected baseline clinical, PB cfDNA, and BM plasma cfDNA features and progression-free survival. Points show hazard ratios and horizontal bars show 95% confidence intervals on a log scale. Effect units are binary presence versus absence, per feature, per lesion, per ISS stage, or per standard deviation, as specified in the source data. **D,** Harrell concordance indices for ISS-only, cfDNA burden-only, and combined ISS-plus-cfDNA burden Cox models in the complete-case subset with ISS, PB cfDNA burden, and BM plasma cfDNA burden available. The analysis included 36 patients and 17 PFS events. P values for combined models are from likelihood-ratio tests against the ISS-only model. **E,** Baseline PB cfDNA and BM plasma cfDNA feature burden across ISS stage in TFRIM4 patients with available ISS and cfDNA burden data. The analysis included 36 patients: ISS I, n = 10; ISS II, n = 15; ISS III, n = 11. P values are from Kruskal-Wallis tests across ISS stage. These analyses were exploratory and were not powered to establish independent prognostic value.

### Serial cfDNA profiling captures patterns of molecular persistence, reappearance, and clonal evolution

We next examined serial cfDNA profiles to characterize patterns of molecular persistence, reappearance, and clonal evolution across PB and BM compartments in individual patients. These analyses were exploratory and feature-specific, with row-specific denominators reflecting differences in assay availability, sampling density, and BCR callability (Fig. 6). Progressor-oriented summaries restricted to samples within 180 days before progression were small and were interpreted as descriptive case examples rather than estimates of monitoring sensitivity or lead time.

**Figure 6.**
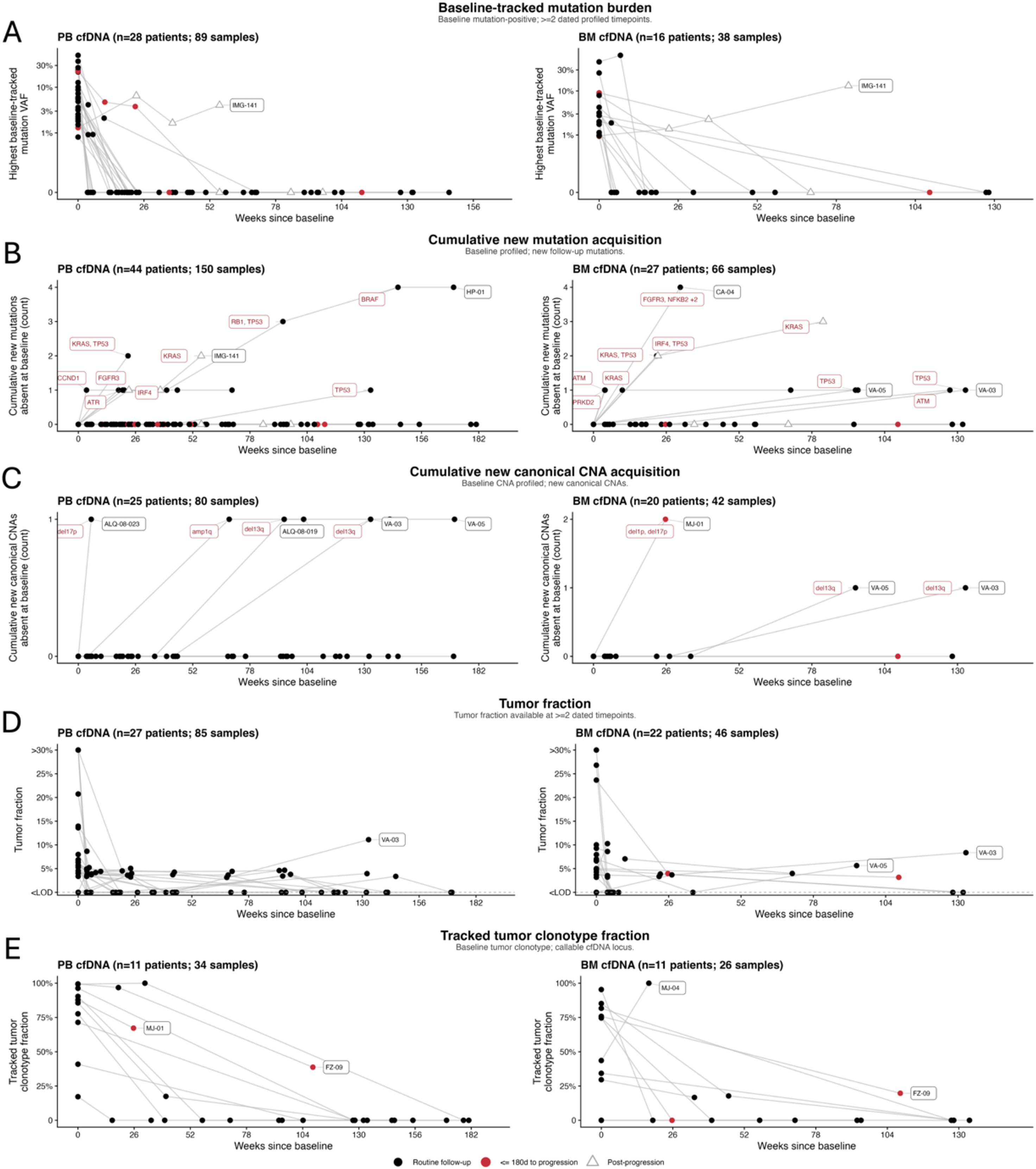
Serial cfDNA profiles capture patterns of molecular persistence, reappearance, and clonal evolution across PB and BM plasma cfDNA compartments. **A**, Highest baseline-tracked mutation VAF over time in PB cfDNA and BM plasma cfDNA. **B,** Cumulative emergent mutations absent from baseline. **C,** Cumulative new canonical copy-number alterations absent from baseline. **D,** Tumor fraction over time. **E,** Tracked baseline BM-cell clonotype fraction over time. For each readout, PB cfDNA is shown on the left and BM plasma cfDNA on the right. Points are plotted by weeks from baseline, and lines connect serial samples from the same patient within each compartment and readout. Symbols indicate routine follow-up, relapse-proximal pre-progression samples collected within 180 days before recorded progression, and post-progression samples. Panel titles report the number of patients and samples evaluable for each compartment-specific analysis. Baseline-tracked mutation and clonotype analyses required a baseline mutation or BM-cell clonotype reference, whereas emergent mutation and CNA analyses required an evaluable baseline assay establishing absence at baseline. Missing assays were retained as missing rather than coded as zero, and denominators differ across panels because molecular readouts have distinct assay availability, sampling density, and callability requirements. Together, these longitudinal profiles show that PB cfDNA and BM plasma cfDNA can capture treatment-associated molecular response, residual or recurrent tumor burden, and emergent genomic lesions over time, while requiring feature-class-specific interpretation across compartments.

Across serial samples, baseline-tracked mutation burden (Fig. 6A), emergent mutation acquisition (Fig. 6B), new canonical CNA acquisition (Fig. 6C), tumor fraction (Fig. 6D), and tracked BCR clonotype fraction (Fig. 6E) provided complementary readouts of persistence, reappearance, and molecular evolution. Emergent mutations and newly appearing canonical CNAs included events involving *TP53*, *KRAS*, *BRAF*, *FGFR3*, *ATM*, *RB1*, *CCND1*, *IRF4*, *PRKD2*, and *NFKB2*, and new del13q, del17p, del1p, and amp1q (Fig. 6B,C). Non-progressors more often showed low or intermittent molecular signal, although a persistent signal without recorded progression occurred in a subset, particularly in PB cfDNA. Overall, these serial profiles capture distinct patterns of clonal dynamics, including persistent tumor signal through treatment, reappearing mutations at disease evolution, and emergent genomic lesions, but do not establish clinical sensitivity, lead time, or monitoring performance in the absence of prospective validation with defined sampling intervals and clinical endpoints (Table 3). Our findings highlight that while cfDNA is a versatile tool for longitudinal monitoring, its clinical interpretation must account for tumor fraction, assay detection limits, and the specific molecular compartment being sampled.

**Table 3.**
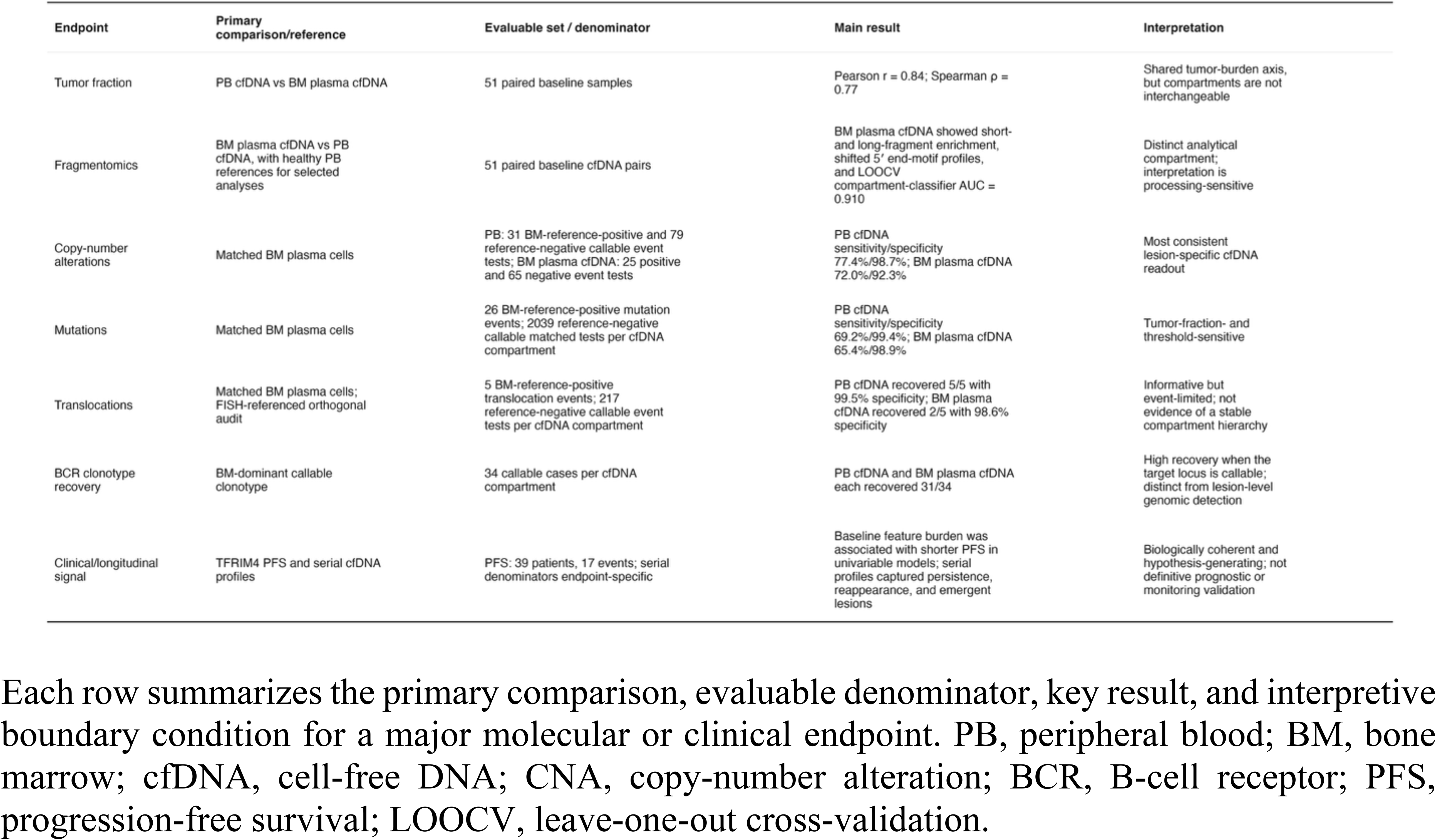
Endpoint-specific interpretation framework for matched myeloma cfDNA profiling.

## Discussion

In this matched cross-compartment study, we show that liquid-biopsy performance in MM cannot be summarized by a single cfDNA-versus-marrow concordance measure. PB cfDNA, BM plasma cfDNA, and BM plasma cells provided complementary but non-equivalent views of disease, and their relationships depended on sampling compartment, molecular feature class, tumor fraction, assay callability, threshold selection, processing context, and reference-standard uncertainty. CNAs were the most consistent lesion-specific cfDNA readout, mutation recovery was threshold- and burden-sensitive, translocation estimates were informative but event-limited, BCR clonotype recovery was high when the dominant clonotype was callable, and fragmentomic differences distinguished BM plasma cfDNA from PB cfDNA while remaining sensitive to pre-analytical handling (Table 3). These findings argue against a single cfDNA-versus-marrow concordance metric and support compartment-aware interpretation of myeloma liquid biopsies.

Prior studies have shown that PB cfDNA can recover clinically relevant myeloma alterations and provide longitudinal readouts of treatment response and clonal evolution ^13–17,26,27^. We extend these findings by directly comparing BM plasma cells, BM plasma cfDNA, and PB cfDNA within a matched cohort and by interpreting concordance separately for each endpoint. This distinction matters because aggregate concordance can obscure why signals agree or diverge: CNAs, point mutations, IGH translocations, BCR clonotypes, tumor-fraction estimates, and fragmentomic features differ in biological meaning, assay callability, quantitative scale, and threshold sensitivity. Under the calibrated framework, CNAs were the most consistent lesion-specific cfDNA readout, whereas mutation recovery scaled with tumor fraction and was more sensitive to threshold selection. cfDNA-only mutations could not be equated with absence from marrow, since a subset showed subthreshold support in matched BM tumor cells below the primary cutoff, and germline and clonal hematopoiesis variants present in buffy-coat DNA were removed by matched-normal calling. These observations support a continuum model in which cfDNA-private calls may reflect low-VAF marrow signal, spatial heterogeneity, or technical noise rather than a binary class of true tumor signal or artifact.

The fragmentomic analyses provide an independent line of evidence that BM plasma cfDNA and PB cfDNA should not be treated as interchangeable liquid compartments. BM plasma cfDNA showed broader fragment-size remodeling, attenuation of the mono-nucleosomal peak, enrichment across both short and long fragment ranges, and directional shifts in 5′ end-motif composition. Persistence of these features at low tumor fraction argues against a purely tumor-derived short-fragment effect, while depletion of CC-starting motifs is consistent with altered nuclease-associated fragmentation that is not explained solely by tumor fraction ^28,29^. At the same time, the larger BM-versus-PB separation in shipped relative to locally processed samples implicates pre-analytical handling, including processing delay, ex vivo leukocyte lysis, dilution by longer genomic DNA, and library-quality differences, as potential contributors to the observed architecture. Thus, BM plasma cfDNA should not be interpreted as having a single intrinsic fragmentomic signature that can be cleanly attributed to marrow biology. Instead, BM plasma cfDNA is a distinct analytical compartment whose signal reflects both biological and pre-analytical factors that must be controlled before fragmentomic interpretation can be generalized ^22–25^. This processing sensitivity defines a key boundary condition for using marrow plasma cfDNA as a fragmentomic analyte.

The clinical and longitudinal analyses provide biologically coherent but exploratory evidence for potential clinical relevance. Continuous baseline cfDNA feature burden was associated with shorter PFS in TFRIM4 in both compartments, cfDNA signal burden increased across ISS stage, and serial profiling captured persistence, reappearance, and emergent molecular signals. These observations align with prior reports linking cfDNA detectability and persistence to disease burden and prognosis in myeloma^13,17,20,27,30,31^. However, modest event counts, endpoint-specific denominators, and variable sampling density preclude claims of independent prognostic validity, monitoring sensitivity, lead time, or clinical actionability. These data support prospective validation but do not define a monitoring standard.

Several limitations apply. The BM plasma-cell reference is clinically grounded but biologically incomplete because it relies on a single-site CD138+ specimen in a spatially heterogeneous disease ^6,10^. Cohorts spanned newly diagnosed and relapsed/refractory disease and contributed unequally to each endpoint, with denominators reported throughout. BM plasma cfDNA was shipped overnight for two of three contributing cohorts, confounding compartment from pre-analytical effects in the fragmentomic analysis. Future studies prioritizing marrow-derived fragmentomics should use immediate stabilization or local processing to decouple biological signal from processing-associated artifacts. Translocation recovery and FISH benchmarking were event-limited. Survival and longitudinal analyses were exploratory and underpowered for monitoring claims. Healthy reference samples were peripheral blood only; a healthy BM plasma cfDNA reference was not available.

Together, these findings support myeloma cfDNA as a calibrated, compartment-aware molecular readout rather than a generic surrogate for marrow. Its interpretation should be structured around feature class, tumor fraction, sampling compartment, assay threshold, and reference definition, rather than a single aggregate concordance metric. PB cfDNA is the more practical substrate for serial genomic monitoring when tumor fraction and assay callability are adequate. BM plasma cfDNA is a marrow-adjacent liquid compartment with distinct fragmentomic behavior, but its interpretation requires strict control of collection and processing conditions, and it did not provide a uniformly higher-yield tumor readout. Prospective validation in uniformly processed cohorts will be needed to establish clinical utility and clarify the biological and pre-analytical drivers of compartment-specific cfDNA architecture.

## Methods

### Study design and analysis cohorts

This study integrated three matched specimen compartments from patients with plasma-cell disorders: CD138-enriched bone marrow plasma cells, BM plasma cfDNA, and PB cfDNA. Samples were obtained from three cohorts. TFRIM4 is a prospective multicentre cohort of patients with newly diagnosed multiple myeloma sampled longitudinally across treatment milestones (diagnosis, post-induction, post-transplant, during maintenance, and at relapse) across 11 Canadian sites. IMMAGINE is a Princess Margaret Cancer Centre cohort enrolling patients with newly diagnosed and relapsed/refractory plasma-cell disorders with paired peripheral blood and marrow sampling. ALGONQUIN is a multicentre, open-label, phase 1/2 Myeloma Canada Research Network trial evaluating belantamab mafodotin in combination with pomalidomide and dexamethasone in relapsed/refractory multiple myeloma^32^. CD138-enriched BM-cell DNA was available for TFRIM4 and IMMAGINE but not for ALGONQUIN.

The study inventory included 155 patients (95 TFRIM4, 21 IMMAGINE, 39 ALGONQUIN). The primary baseline paired-cfDNA cohort included 74 patients with matched PB and BM plasma cfDNA data from at least one assay (42 TFRIM4, 11 IMMAGINE, 21 ALGONQUIN). Sample availability and assay-level attrition are summarized in Extended Data Fig. 1A. Endpoint-specific analyses used narrower evaluable populations defined by assay availability, matched reference data, and locus- or event-level callability. For BM plasma-cell–referenced genomic recovery, the reference compartment was the matched baseline CD138-enriched BM plasma-cell specimen from the same patient. Longitudinal and survival analyses used patients with required dated samples and clinical annotations, with denominators reported separately.

### Sample processing and DNA extraction

Peripheral blood was collected in Streck Cell-Free DNA BCT CE tubes (cat. #218997; ≥15 ml target) and shipped overnight from collection sites at ambient temperature to the Pugh laboratory at Princess Margaret Cancer Centre. Plasma was separated by centrifugation at 1,900 × g for 10 min at 4 °C. Following the initial centrifugation, the plasma fraction was transferred to a fresh tube for a second centrifugation at 16,000 × g for 10 min at 4 °C, while the buffy coat was retained for genomic DNA extraction. Clarified plasma and buffy coat were stored at −80 °C. Buffy-coat genomic DNA was extracted with the QIAGEN AllPrep DNA/RNA/miRNA Universal Kit (cat. #80224); plasma cfDNA with the QIAGEN QIAamp Circulating Nucleic Acid Kit (cat. #55114).

BM aspirate processing differed by cohort and collection site. TFRIM4 BM aspirates were collected in EDTA tubes and shipped to the Reiman laboratory at Dalhousie University in Saint John, New Brunswick, for CD138+ enrichment using the EasySep Human CD138 Positive Selection Kit II (STEMCELL Technologies, cat. #17877). Genomic DNA was extracted with the QIAGEN DNeasy Blood and Tissue Kit (cat. #69506); yield/quality assessed by Qubit 3.0 Fluorometer (Thermo Fisher) and the Agilent 2200 TapeStation System. DNA and BM plasma were then shipped to the Pugh laboratory in Toronto for library preparation and sequencing. IMMAGINE BM samples were collected and processed locally at Princess Margaret Cancer Centre. ALGONQUIN was a multicentre trial in which BM samples from participating sites (n = 6) were sent to Princess Margaret Cancer Centre for processing; samples collected at the Toronto/Princess Margaret site were therefore locally processed, whereas samples from non-Toronto ALGONQUIN sites were shipped to Toronto before processing. IMMAGINE and ALGONQUIN BM samples were processed using the same enrichment workflow as TFRIM4, with DNA extraction by AllPrep (cat. #80224). All available BM plasma supernatant was processed for cfDNA extraction at the Pugh laboratory using QIAGEN QIAamp Circulating Nucleic Acid Kit (cat. #55114); BM CD138+ cells were not routinely available for ALGONQUIN, reflected in Extended Data Fig. 1A.

### Targeted-panel design, library preparation, and sequencing

Targeted libraries were prepared from PB cfDNA, BM plasma cfDNA, CD138-enriched BM-cell DNA, and matched normal DNA using the KAPA HyperPrep Kit (Roche; cat. #07962363001). CD138-enriched BM-cell DNA, and matched normal DNA were fragmented to 250bp using the Covaris LE220 ultrasonicator. cfDNA libraries used IDT xGen Dual Index UDI CS Adapters (cat. #1080799) and custom IDT xGen Lockdown hybrid-capture probe pools (cat. #1072281). cfDNA libraries used a minimum input of 20 ng where available; capture pooled 2–4 libraries (∼150–500 ng each), followed by AMPure XP cleanup and Qubit quantification. Cohort- and sample-type-specific assay implementation (panel components, sequencing platform, AMDL pipeline use, BM CD138+ pre-2023 Bratman barcode handling (Newman et al., 2014) is detailed in Supplementary Methods.

The custom capture design spanned 257,803 bp and included two components: (i) 37 recurrently altered myeloma genes (prognostic, IMiD-resistance, therapeutic-pathway; 136,588 bp) and (ii) canonical myeloma translocation hotspots (IGH/MYC partners including *FGFR3/NSD2*, *CCND1*, *MAF*, *MAFB*, *CCND3*, *MYC*) and immunoglobulin V(D)J regions (*IGH*, *IGK*, *IGL*; 121,215 bp) (Extended Data Fig. 1C). The full target list is provided in Supplementary Table 13.

Libraries were sequenced at the Princess Margaret Genomics Centre using paired-end 2 × 150 bp Illumina sequencing (NextSeq 550 for gene-panel libraries to maintain consistency with AMDL; NovaSeq 6000 for immunoglobulin/translocation libraries). Targeted sequencing aimed for 10,000× raw on-target mean coverage for cfDNA samples and ≥1,000× for BM-cell and matched-normal DNA. A subset also underwent shallow whole-genome sequencing (1× mean target) on NovaSeq 6000 for tumor fraction, copy-number, and fragmentomics.

### Read processing, alignment, and consensus generation

Targeted sequencing FASTQs were processed using ConsensusCruncher^33^. Reads were aligned to GRCh38 with BWA-MEM v0.7.15. Read groups were harmonized with Picard v2.10.9; BAMs sorted with samtools v1.20. Targeted BAMs underwent local indel realignment with GATK v3.3-0 before UMI-aware consensus generation. Mutation, translocation, and immunoglobulin analyses used these consensus BAMs. sWGS FASTQs were aligned with BWA-MEM v0.7.15 without consensus collapsing; duplicates were marked with Picard v2.10.9 and BAMs indexed with samtools v1.20.

### Somatic mutation calling and threshold calibration

Somatic short variants were called against matched normal DNA using an ensemble (MuTect v1.1.5, MuTect2/GATK v3.8, Strelka v2.9.10, SomaticSniper v1.0.5.0, VarDict v1.7.0) implemented through the Pugh Lab pipeline suite v0.9.5 (github.com/pughlab/pipeline-suite).

Calling against a matched buffy-coat normal sample was used to remove germline variants and clonal hematopoiesis variants detectable in the matched leukocyte DNA. Calls required a caller pass-like filter status, ≥3 alternate reads, and VAF ≥ 0.0005, and were retained if called by MuTect2 or by ≥2 additional callers.

Variants were annotated with Ensembl VEP v98, vcf2maf v1.6.17, ExAC/gnomAD, and OncoKB (tumor type: plasma cell myeloma)^34–37^. Variants were restricted to capture space (25 bp probe padding) and filtered using population-frequency thresholds and ENCODE blacklist overlap^38^. Variants exceeding population-frequency filters (PopAF_max <= 0.001) were excluded unless annotated as oncogenic or likely oncogenic by OncoKB.

The BM plasma-cell mutation reference required VAF ≥ 0.10 and ≥ 4 alternate reads, defining a high-confidence clonal anchor for benchmarking rather than a full low-VAF marrow catalogue. The primary cfDNA Tier 1 threshold (VAF ≥ 0.0075, ALT ≥ 12, depth ≥ 500) was selected to balance BM-referenced balanced accuracy, driver sensitivity, and cfDNA-only event burden. A lower-support exploratory Tier 2 (VAF ≥ 0.005, ALT ≥ 8, depth ≥ 250) was retained for sensitivity analyses but not used for primary concordance, detectability, or clinical claims (Supplementary Methods; Extended Data Fig. 4; Supplementary Tables 3–4). After threshold selection, recurrent low-confidence non-oncogenic cfDNA-only patterns were demoted from Tier 1 to Tier 2, and residual Tier 1 cfDNA-only mutations were reviewed against matched BM-cell data to distinguish subthreshold marrow support, plausible spatial/compartmental heterogeneity, and likely technical noise (Supplementary Methods).

### Translocation calling and framework selection

Canonical myeloma translocations were called from targeted-panel data using IgCaller v1.4-beta, filtered for caller support, blacklist overlap, immunoglobulin-side plausibility, and proximity to canonical partner loci (*CCND1*, *NSD2/FGFR3*, *MAF*, *MAFB*, *CCND3*, *MYC*). The selected primary framework (tx_shared_cf100_rescue400) used compartment-specific IgCaller settings and calibrated score thresholds: BM cell calls required clean canonical support with score ≥ 40; cfDNA compartments required clean canonical support with score ≥ 100; high-confidence clean near-canonical events were rescued at score ≥ 400. cfDNA and BM-cell scores are directly comparable because IgCaller was run with lower expected tumor purity for cfDNA (5%) than for BM cells (60%) (Supplementary Methods).

### CNA calling and tumor fraction

sWGS BAMs were processed with ichorCNA v0.3.2 to estimate tumor fraction and call recurrent myeloma CNAs^39^. Read counts were summarized in 1 Mb bins (readCounter, hmmcopy_utils 170718; MAPQ ≥ 20), GC- and mappability-corrected, and normalized against the hg38 ultra-low-pass panel of normals. ichorCNA was run on GRCh38 with candidate ploidies 2 and 3, normal-fraction grid 0.50–0.95, max copy number 5, autosomes for training, subclonal state estimation, and minimum segment size 50 bins. Samples with ≥ 5% tumor fraction by ichorCNA were considered high tumor fraction.

Recurrent arm-level CNAs (del1p, amp1q, del13q, del17p) were called when ≥ 40% of the relevant chromosome arm was callable and ≥ 50% of the callable arm length showed the expected altered state. Hyperdiploidy-related gains were summarized across chromosomes 3, 5, 7, 9, 11, 15, 19, and 21 (≥ 70% of length showing gain); relaxed hyperdiploidy required gain of ≥ 5 of these 8 chromosomes. The selected primary framework (cna_shared_alt0.5_cov0.4_tf0) applied uniform thresholds across compartments and did not impose a hard tumor-fraction mask. Samples with TF < 3% were flagged as low-purity for categorical interpretation but were not masked under the primary framework (Supplementary Methods).

### BCR clonotype recovery

Immunoglobulin clonotypes were inferred from targeted-panel FASTQs using MiXCR v3.0.13 under Java 8 (RNA-seq preset; species hsa; partial-alignment rescue; VGeneWithP). Two rounds of partial-assembly rescue were followed by extension and clonotype assembly; clones were exported per locus (IGH, IGK, IGL). Productive clonotypes had non-missing CDR3 amino-acid sequences without stop codons or frameshift markers. A clonotype was defined by locus, CDR3 amino-acid sequence, and top V/J calls.

A BM plasma-cell clonotype qualified as the dominant baseline reference if its top productive locus clonotype had ≥ 5 reads, ≥ 10% of productive reads, no top-read tie, and ≥ 1.5× dominance over the runner-up when present. In the cfDNA target compartment, the matched locus was callable with ≥ 10 productive reads, and the BM-matched clonotype was considered recovered when ≥ 5 target reads matched the BM-dominant clonotype (Supplementary Methods).

### Fragmentomics analysis

Fragmentomics analyses used cfDNA sWGS data via the Pugh lab fragmentomics pipeline v0.9.11 (<u>github.com/pughlab/pipeline-suite</u>) with samtools v1.20, Picard v2.10.9, sambamba v0.7.0, bedtools v2.27.1, Python v3.7.2, R v4.1.0, and Griffin v0.1.0. We evaluated short-fragment fraction (<150 bp), median insert size, insert-size distribution shape, 5′ end-motif summaries, and DELFI-like regional-fragmentation ratios restricted to 5-Mb bins overlapping recurrent myeloma loci (1p22/*CDKN2C*, 1q21/*CKS1B*, 4p16/*FGFR3-MMSET*, 8q24/*MYC*, 11q13/*CCND1*, 13q14/*RB1*, 14q32/*IGH*, 16q23/*MAF*, 17p13/*TP53*, 20q12/*MAFB*). Locally processed healthy PB controls (n = 26) from the CHARM Consortium served as supplementary references. Reference distances were computed from sample-by-feature matrices as deviations from the relevant centroid. For 5′ end motifs, distance to the healthy PB centroid was defined as the mean absolute deviation of each sample’s motif-frequency profile from the mean motif-frequency profile of healthy PB controls. PCA was performed on centered and scaled insert-size profiles, 5′ end-motif frequencies, and DELFI-like 5-Mb regional fragmentation ratios, and was used descriptively to visualize compartment structure and proximity to healthy PB and low-TF cfDNA reference groups.

To quantify compartment-level fragmentomic separability, we trained logistic-regression classifiers to distinguish baseline BM plasma cfDNA from matched baseline PB cfDNA using eight fragmentomic summary features: median insert size, short-fragment fraction, long-fragment fraction, mono-/di-nucleosomal fragment ratio, insert-size entropy, mean absolute 5-Mb fragmentation-ratio signal across myeloma-relevant loci, 5′ end-motif entropy, and 5′ end-motif distance to a healthy PB cfDNA centroid. Analyses were restricted to patients with baseline cfDNA available from both compartments. Classifier performance was evaluated using leave-one-patient-out cross-validation (LOOCV), in which all cfDNA samples from the held-out patient were excluded from model training to prevent patient-level leakage between paired PB and BM plasma cfDNA specimens. Cross-validated predicted probabilities for the BM plasma cfDNA class were pooled across held-out folds to generate receiver-operating-characteristic curves and AUC estimates, with BM plasma cfDNA specified as the positive class. Statistical significance for the primary AUC was assessed by within-patient permutation of compartment labels, preserving patient structure while permuting PB/BM plasma cfDNA labels within each patient (Supplementary Methods).

### Statistical analysis

Statistical analyses were performed in R 4.5.2. Continuous variables were summarized using medians and interquartile ranges. Pearson correlation was used for approximately linear relationships and Spearman rank correlation for monotonic relationships or non-normal data. Matched PB cfDNA and BM plasma cfDNA comparisons used two-sided Wilcoxon signed-rank tests; unpaired group comparisons used Wilcoxon rank-sum tests. Fisher’s exact tests were used for categorical comparisons. Sensitivity, specificity, detection rates, and related binary concordance metrics were calculated from prespecified 2 × 2 event counts; 95% confidence intervals were calculated using Wilson score intervals.

Survival analyses were exploratory. Kaplan–Meier curves were compared using log-rank tests; Cox proportional-hazards models estimated hazard ratios with 95% CIs. Most Cox analyses were univariable; selected multivariable models adjusted for ISS stage, PB cfDNA and BM plasma cfDNA feature burden, IMWG cytogenetic risk, and study-stratified baseline hazards. Nested Cox models were compared using likelihood-ratio tests. Unless otherwise specified, tests were two-sided with nominal P < 0.05. Feature-screening analyses were adjusted using the Benjamini– Hochberg method. Analyses with limited sample size, sparse events, or non-estimability were interpreted descriptively.

### Data availability

Sequencing data generated in this study contain human genomic information and are being deposited in EGA for controlled access. Requests will be reviewed according to the University Health Network Data Access Committee, subject to participant consent and institutional data-sharing restrictions. De-identified clinical and sample metadata required to reproduce the reported analyses will be made available through the controlled-access record where permitted. Source data underlying the main and Extended Data Figures are provided with this paper.

### Code availability

Code required to reproduce the main analyses and figures is available at https://github.com/pughlab/cfdna-myeloma-multiomics/. The repository includes the analysis scripts, shared helper functions, script inventory, rerun order, environment setup, and software version documentation. An archived release with a DOI will be provided before publication.

### Ethics approval

All studies were conducted in accordance with the Declaration of Helsinki and were approved by the relevant institutional research ethics boards. TFRIM4 (protocol #17-5429), IMMAGINE (protocol #16-5260), ALGONQUIN (protocol #18-5911), and CHARM (protocol #19-6239) received ethics approval at participating institutions. All participants provided written informed consent for the collection, genomic profiling, and research use of peripheral blood, bone marrow, and associated clinical data. Clinical and sequencing data were analyzed using de-identified study identifiers.

## Acknowledgements

We gratefully acknowledge all study participants and the clinical, research, laboratory and data-coordination teams who contributed to this work. We thank members of the Pugh and Trudel laboratories for ongoing discussions and support. We also acknowledge the contributions of the Princess Margaret Genomics Centre staff and the UHN High-Performance Computing and Bioinformatics Core.

## Author Contributions

D.D.A. led the study, with supervision from S.T. and T.J.P. D.D.A. developed the analytical strategy, selected and prioritized samples for sequencing, coordinated sample organization and submission, performed the computational and statistical analyses, interpreted the results, generated the figures and tables, and drafted and revised the manuscript with support from S.T. and T.J.P. J.E. led biospecimen processing, sequencing coordination, and wet-laboratory sample tracking, with support from S.P. and E.N.W. A.W. and S.S. assisted with bone marrow processing and bone marrow cell DNA extraction for TFRIM4, with study coordination and clinical input from A.R. D.S.S. assisted with clinical data retrieval and sample annotation. A.D., J.P.B., and S.D.P. supported the development of computational analyses and analytical methods. D.W., I.S., K.S., and A.R. enrolled patients and provided biospecimens and clinical data. A.R. led the TFRIM4 study with support from A.M., S.T., and T.J.P. conceived the study, supervised the research, secured funding, and guided study design, data interpretation, and manuscript preparation. All authors reviewed and approved the final manuscript.

## Extended Data Figures

**Extended Data Figure 1.**
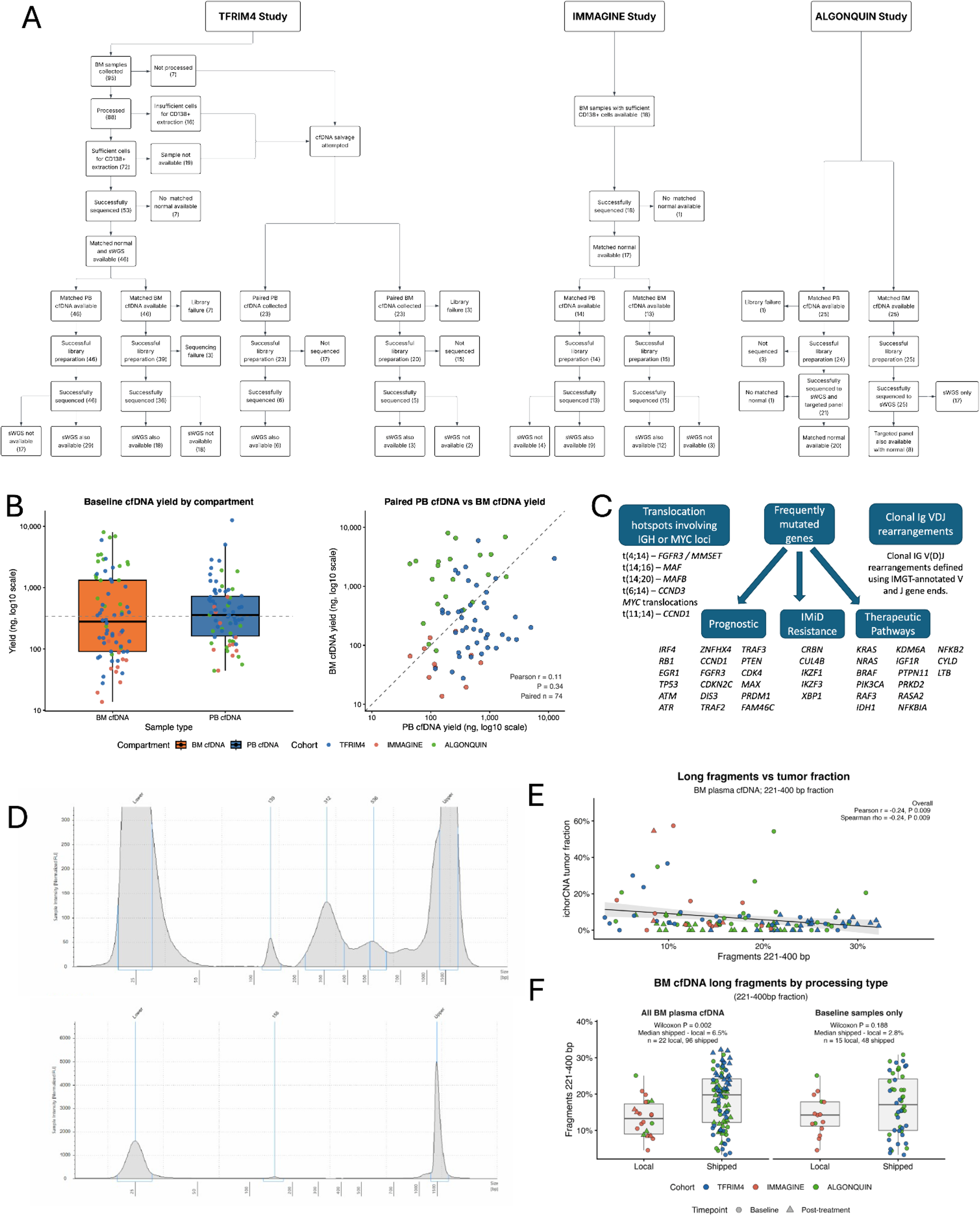
Cohort-specific sample flow, assay availability, cfDNA yield, targeted-panel design, and BM plasma cfDNA processing context. **A**, Cohort-specific sample flow diagrams for TFRIM4, IMMAGINE, and ALGONQUIN, showing how collected baseline specimens contributed to BM plasma-cell, PB cfDNA, and BM plasma cfDNA analysis populations. **B,** Baseline cfDNA yield by compartment and paired PB versus BM plasma cfDNA yield among patients with both compartments available; the diagonal indicates equal yield. **C,** Targeted-panel content used for mutation, translocation, and BCR analyses, including recurrent IGH/MYC translocation loci, myeloma-relevant genes, and clonal IG V(D)J regions. **D,** Representative electrophoretic BM plasma cfDNA profiles showing successful cfDNA-like libraries and failed libraries with prominent adapter signal and little or no sample peak, consistent with low recoverable input and/or dilution by longer-fragment genomic DNA. **E,** Association between BM plasma cfDNA long-fragment fraction, defined as 221–400 bp fragments, and ichorCNA-inferred tumor fraction. The fitted trend shows a weak inverse association, consistent with long-fragment enrichment partially diluting BM plasma cfDNA tumor-fraction estimates. **F,** BM plasma cfDNA long-fragment fraction by processing type for all samples and for baseline samples only. Shipped samples showed higher 221–400 bp fractions than locally processed samples across all timepoints, while the baseline-only comparison showed the same direction but was not statistically significant. In **B, E,** and **F,** points are colored by cohort; in **E** and **F**, point shapes indicate timepoint. Boxplots show median, interquartile range, and 1.5× interquartile range whiskers. Together, these panels define the sample-flow, assay-availability, input-yield, technical-processing, and pre-analytical context underlying the compartment-resolved cfDNA analyses.

**Extended Data Figure 2.**
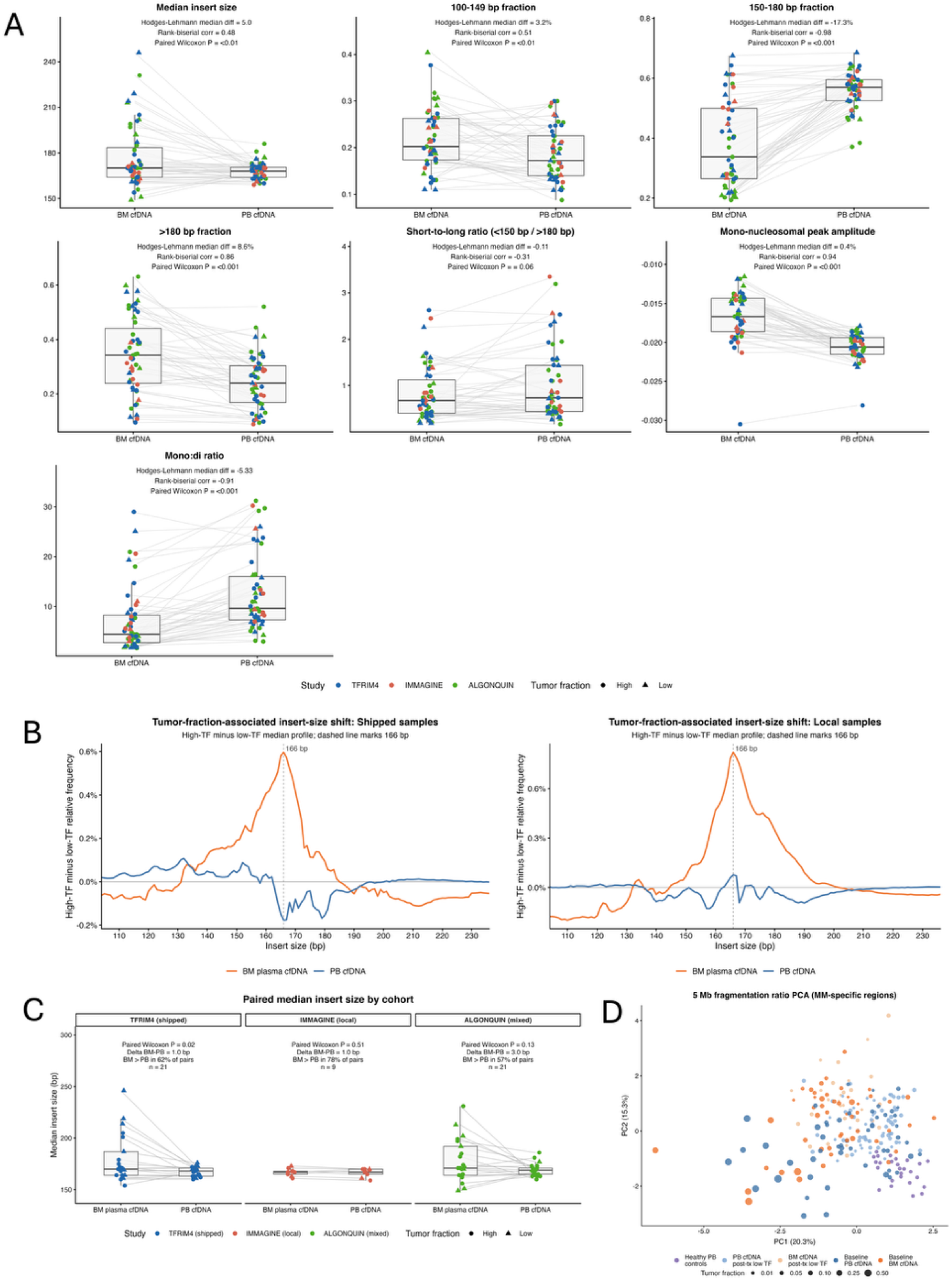
Paired fragment-size, nucleosomal, and regional-fragmentation features in BM plasma cfDNA and PB cfDNA. **A**, Paired baseline fragment-size summary metrics in matched BM plasma cfDNA and PB cfDNA samples. Panels show median insert size, 100-149 bp fraction, 150-180 bp fraction, >180 bp fraction, short-to-long fragment ratio, mono-nucleosomal peak amplitude, and mono:di-nucleosome ratio. Lines connect matched compartments from the same patient; points are colored by cohort and shaped by tumor-fraction category. On-panel labels report paired Hodges-Lehmann median differences, rank-biserial correlations, and paired Wilcoxon test results. **B,** High-minus low-tumor-fraction insert-size difference profiles for BM plasma cfDNA and PB cfDNA, highlighting compartment-specific redistribution around the mono-nucleosomal peak across local and shipped cases. **C,** Paired baseline median insert size stratified by study, with matched BM plasma cfDNA and PB cfDNA samples connected within TFRIM4 (shipped), IMMAGINE (local), and ALGONQUIN (mixed). **D,** Principal-component analysis of 5 Mb regional fragmentation-ratio features across myeloma-relevant genomic regions, including healthy PB controls, post-treatment low-tumor-fraction PB cfDNA and BM plasma cfDNA samples, and baseline PB cfDNA and BM plasma cfDNA samples. Point size denotes tumor fraction for patient-derived samples. Together, these paired and regional-fragmentation analyses show that BM plasma cfDNA and PB cfDNA differ across multiple insert-size and nucleosome-associated features, including enrichment of short and long fragments and altered nucleosomal structure in BM plasma cfDNA, with study-specific variation in median insert-size shifts. These data support the compartment-specific fragmentomic architecture summarized in Figure 2.

**Extended Data Figure 3.**
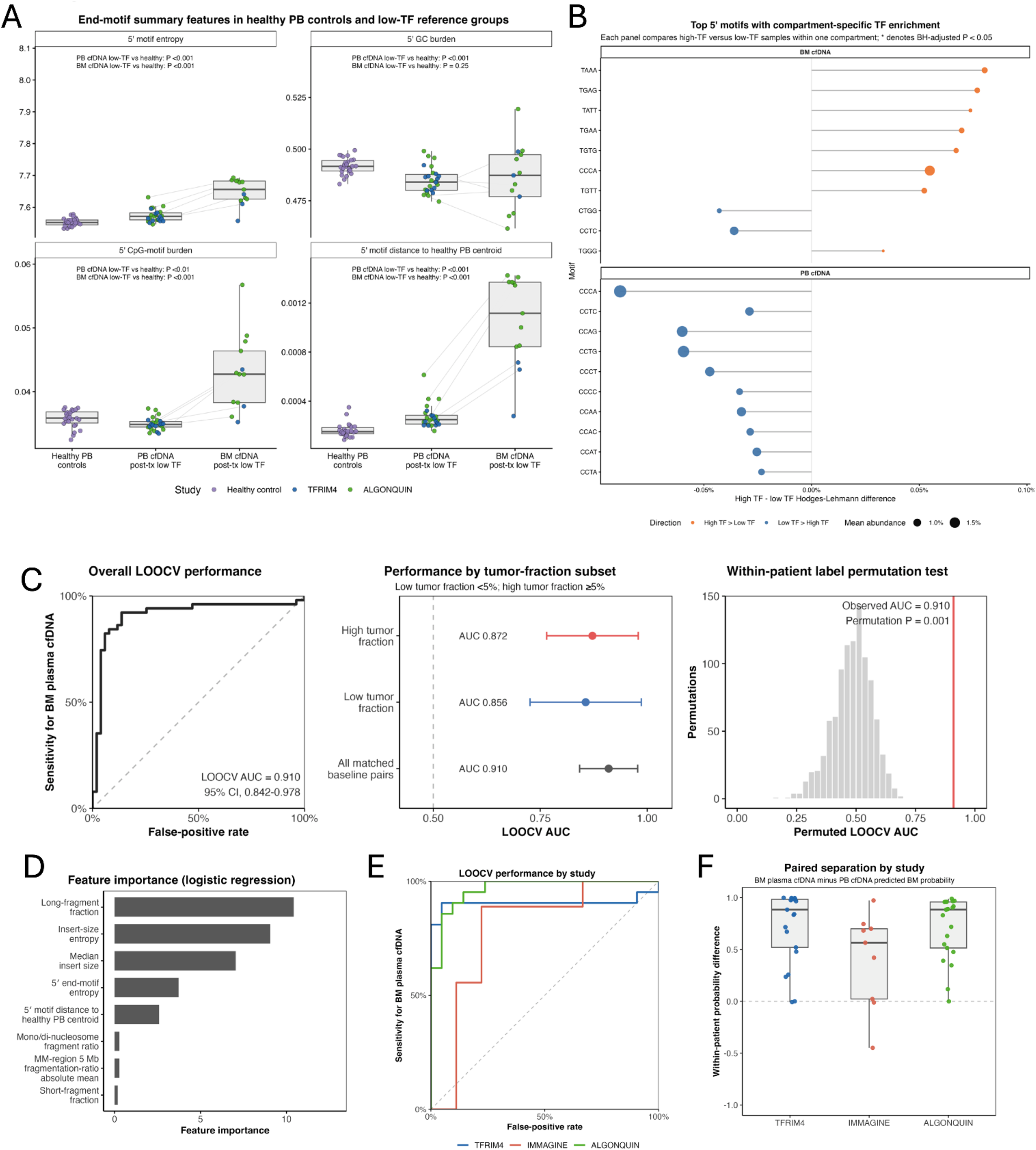
End-motif and fragmentomic features support compartment- and processing-aware interpretation of BM plasma cfDNA and PB cfDNA. **A**, 5′ end-motif summary features in healthy PB controls, low-tumor-fraction post-baseline PB cfDNA, and low-tumor-fraction post-baseline BM plasma cfDNA. Panels show 5′ motif entropy, 5′ GC burden, 5′ CpG-motif burden, and distance from the healthy PB centroid; matched PB/BM plasma cfDNA pairs are connected where available. **B,** Top 5′ end motifs enriched in high-versus low-tumor-fraction samples within each compartment. Hodges-Lehmann differences are shown; point size indicates mean motif abundance, color indicates enrichment direction, and asterisks denote false-discovery-adjusted significance. **C,** Primary all-sample LOOCV logistic-regression classifier using fragment-size and 5′ end-motif summary features to distinguish BM plasma cfDNA from PB cfDNA. Left, ROC curve; middle, AUC estimates with 95% confidence intervals for all pairs and tumor-fraction subsets; right, within-patient compartment-label permutation test, with the observed AUC shown by the vertical line. **D,** Feature importance for the primary classifier, calculated from absolute standardized coefficients. **E**, Study-stratified LOOCV ROC curves for TFRIM4, IMMAGINE, and ALGONQUIN, showing classifier performance within each contributing cohort. **F**, Study-stratified within-patient separation in predicted BM plasma cfDNA probability, calculated as the predicted probability for BM plasma cfDNA minus the predicted probability for matched PB cfDNA within each patient. Positive values indicate higher predicted BM-compartment probability for the BM plasma cfDNA sample than for the matched PB cfDNA sample. Overall, low-tumor-fraction BM plasma cfDNA remained shifted from healthy PB controls, and baseline BM plasma cfDNA remained distinguishable from PB cfDNA across tumor-fraction strata and study contexts.

**Extended Data Figure 4.**
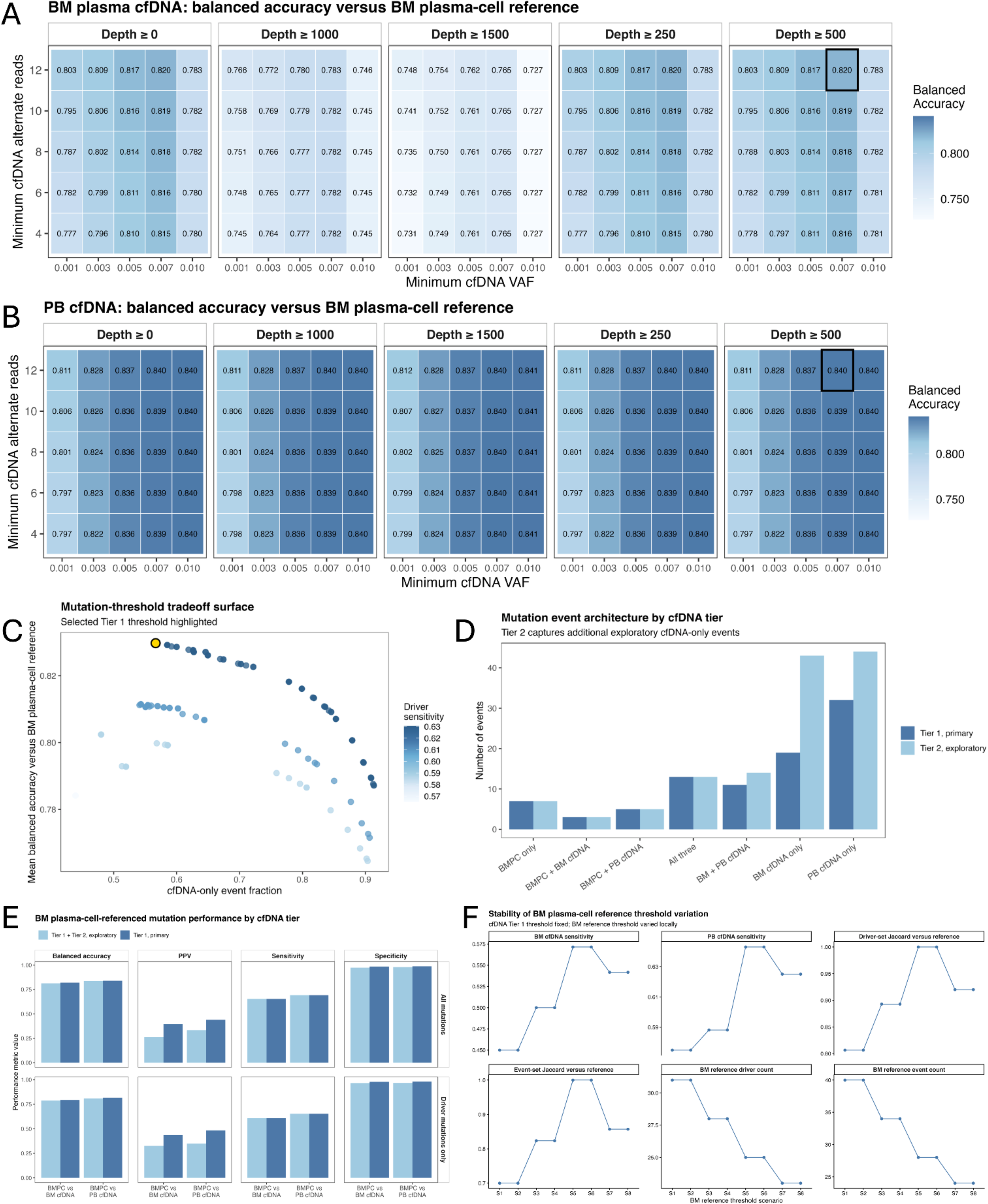
Calibration and stability of BM plasma-cell-referenced cfDNA mutation thresholds. **A**, BM plasma-cell-referenced balanced accuracy for BM plasma cfDNA mutation calling across candidate cfDNA thresholds, varying minimum cfDNA VAF, minimum alternate-read support, and depth filters. The selected primary Tier 1 threshold is highlighted. **B,** Corresponding balanced-accuracy grid for PB cfDNA using the same candidate threshold framework and BM plasma-cell reference. **C,** Mutation-threshold tradeoff surface showing mean balanced accuracy against the BM plasma-cell reference versus the fraction of cfDNA-only events. Point shading indicates driver sensitivity, and the selected Tier 1 threshold is highlighted. **D,** Mutation event architecture under Tier 1 primary and Tier 2 exploratory cfDNA thresholds, classified by detection in BM plasma cells, BM plasma cfDNA, and PB cfDNA. **E,** BM plasma-cell-referenced mutation performance by compartment and cfDNA tier, including balanced accuracy, positive predictive value, sensitivity, and specificity for all mutations and driver-mutation subsets. **F,** Stability analysis of the BM plasma-cell reference under local BM reference-threshold variation while holding the cfDNA Tier 1 threshold fixed, showing effects on BM plasma cfDNA sensitivity, PB cfDNA sensitivity, driver-set Jaccard similarity, event-set Jaccard similarity, BM reference driver count, and BM reference event count. BM reference threshold scenarios were: S1, VAF ≥0.050, ALT ≥4, depth ≥0; S2, VAF ≥0.050, ALT ≥4, depth ≥50; S3, VAF ≥0.075, ALT ≥4, depth ≥0; S4, VAF ≥0.075, ALT ≥4, depth ≥50; S5, VAF ≥0.100, ALT ≥4, depth ≥0, used as the manuscript reference; S6, VAF ≥0.100, ALT ≥4, depth ≥50; S7, VAF ≥0.125, ALT ≥4, depth ≥0; and S8, VAF ≥0.125, ALT ≥4, depth ≥50. Together, these analyses select the primary cfDNA mutation threshold used for BM plasma-cell-referenced concordance analyses, quantify the tradeoff between sensitivity and cfDNA-only calls, and show that apparent cfDNA recovery depends partly on the stringency of the BM plasma-cell reference.

**Extended Data Figure 5.**
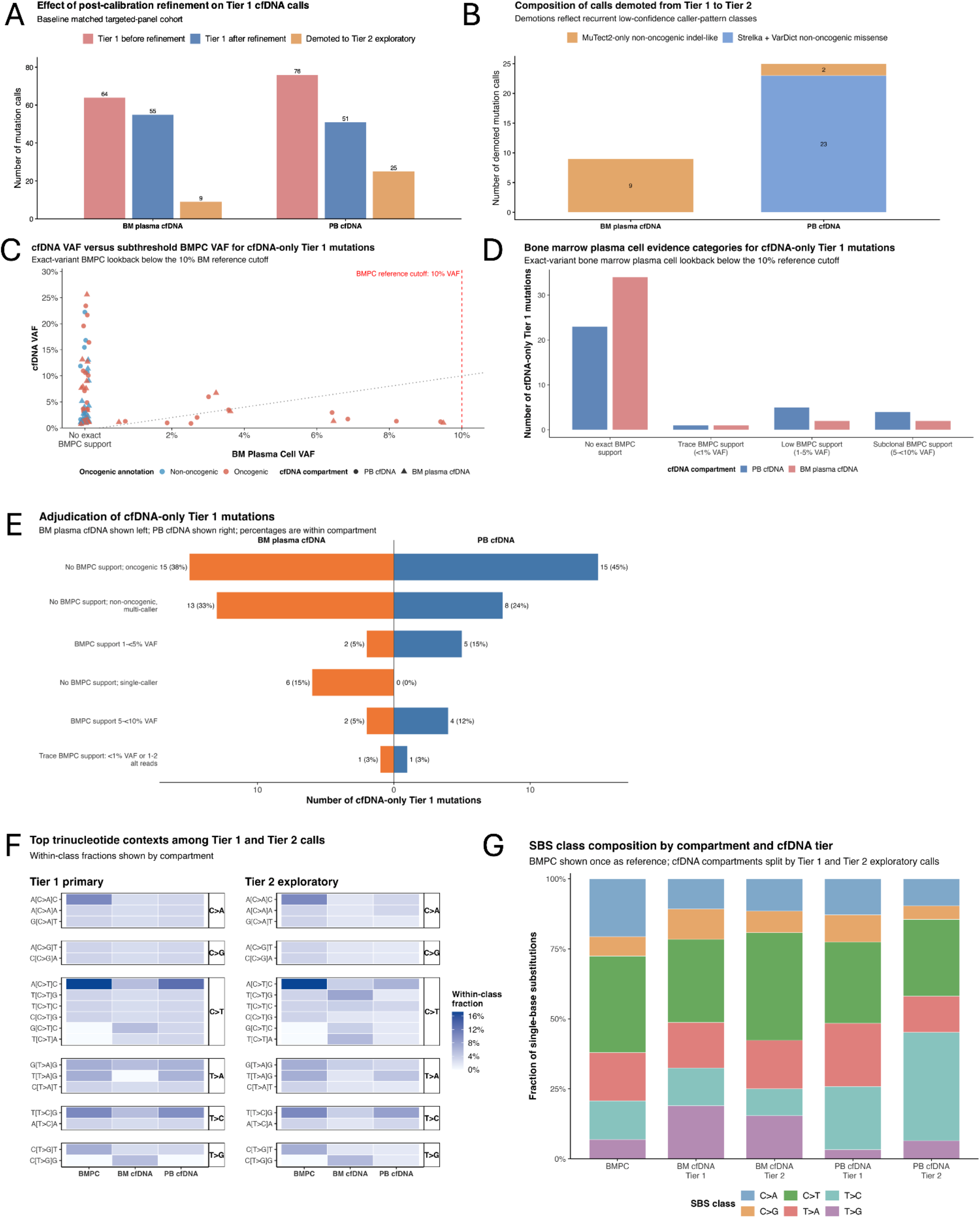
Post-calibration refinement and adjudication of cfDNA-only mutation calls. **A**, Effect of post-calibration refinement on Tier 1 cfDNA mutation calls in the baseline matched targeted-panel cohort, shown separately for BM plasma cfDNA and PB cfDNA. Bars show Tier 1 calls before refinement, Tier 1 calls retained after refinement, and calls demoted to Tier 2 exploratory status. **B,** Composition of calls demoted from Tier 1 to Tier 2, separating recurrent low-confidence caller-pattern classes. **C,** Exact-variant BM plasma-cell lookback for cfDNA-only Tier 1 mutations, comparing cfDNA VAF with subthreshold BM plasma-cell VAF below the 10% BM reference cutoff. Points are colored by cfDNA compartment and annotated by OncoKB oncogenic or likely oncogenic status. **D,** BM plasma-cell evidence categories for cfDNA-only Tier 1 mutations, including no exact BM plasma-cell support, trace support below 1% VAF, low-level support between 1% and 5% VAF, and subclonal support between 5% and 10% VAF. **E,** Adjudication of cfDNA-only Tier 1 mutations by compartment, separating variants with subthreshold BM plasma-cell support, variants without BM support but oncogenic plausibility, non-oncogenic multi-caller support, single-caller support, and residual uncertainty. **F,** Trinucleotide-context composition among Tier 1 primary and Tier 2 exploratory single-nucleotide variant calls across BM plasma cells, BM plasma cfDNA, and PB cfDNA. **G,** SBS-class composition by compartment and cfDNA tier, with BM plasma cells shown as the reference comparator and cfDNA compartments split into Tier 1 primary and Tier 2 exploratory calls. Together, these analyses show that post-calibration refinement demoted recurrent low-confidence cfDNA-only patterns without discarding them, and that residual cfDNA-only Tier 1 mutations are not a single class of false positives. Instead, they include variants with subthreshold BM plasma-cell support, plausible compartment-specific or spatially heterogeneous signal, and unresolved technical uncertainty.

**Extended Data Figure 6.**
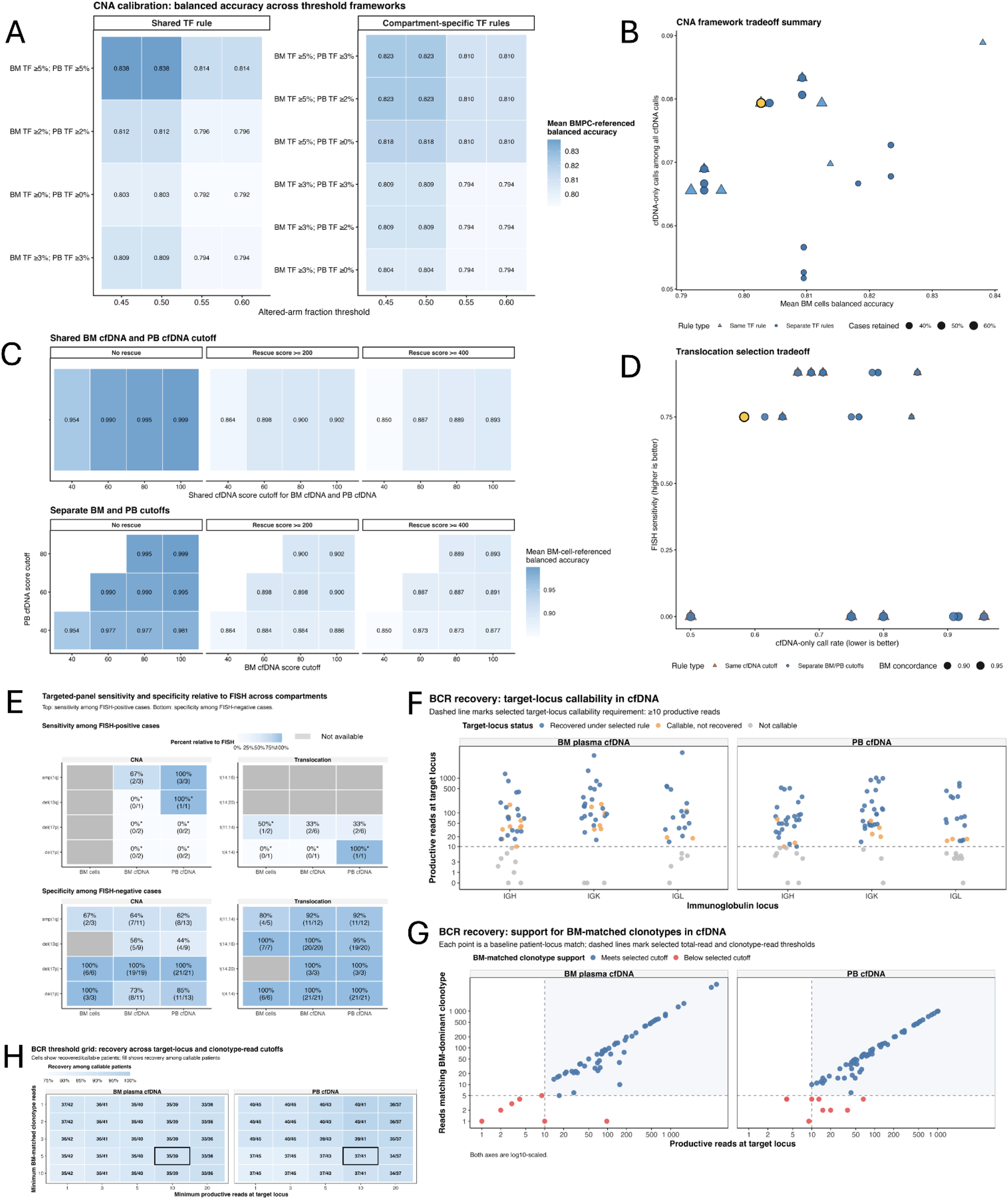
Calibration and benchmarking of CNA, translocation, and BCR recovery in BM plasma cfDNA and PB cfDNA. **A**, Candidate copy-number alteration calibration frameworks evaluated against the BM plasma-cell reference. Heat maps show mean balanced accuracy across altered-arm fraction thresholds and tumor-fraction callability rules, including shared and compartment-specific BM/PB cfDNA requirements. **B,** CNA framework tradeoff summary showing balanced accuracy versus the fraction of cfDNA-only calls among all cfDNA calls, with point annotations indicating rule type and case retention. **C,** Candidate translocation calibration frameworks evaluated across BM and cfDNA score cutoffs, shared versus separate BM/PB cfDNA thresholds, and high-specificity rescue settings. Heat maps show mean BM plasma-cell-referenced balanced accuracy. **D,** Translocation selection tradeoff showing cfDNA-only call rate versus FISH-referenced sensitivity across candidate rule sets. **E,** Orthogonal FISH benchmarking of targeted-panel CNA and translocation calls across BM plasma cells, BM plasma cfDNA, and PB cfDNA. Heat maps show sensitivity among FISH-positive cases and specificity among FISH-negative cases, with percentages and event counts shown in each cell; grey cells indicate unavailable comparisons. **F,** BCR target-locus callability in BM plasma cfDNA and PB cfDNA, showing productive reads at IGH, IGK, and IGL loci relative to the target-locus callability threshold. **G,** BM-matched clonotype support in cfDNA, comparing productive reads at the target locus with reads matching the BM-dominant clonotype; dashed lines mark selected target-read and clonotype-read thresholds. **H,** BCR threshold grid showing recovery of BM-matched clonotypes across target-locus and clonotype-read cutoffs among callable patients. Highlighted cells indicate the selected recovery rule. Together, these analyses define endpoint-specific calibration frameworks for CNA, translocation, and BCR recovery, with calibration summaries provided in Supplementary Tables 10-12.

## Supplementary Table Legends

**Supplementary Table 1. Baseline cohort, assay availability, cfDNA yield, and sample metadata.**

Baseline patient- and sample-level metadata supporting Figure 1 and Extended Data Fig. 1. The canonical paired-cfDNA cohort included 74 patients. cfDNA yield, assay availability, and QC summaries use row-specific denominators according to available compartment, assay, and sample metadata.

**Supplementary Table 2. Paired baseline fragmentomics, tumor-fraction strata, and processing-context sensitivity**

Paired baseline BM plasma cfDNA and PB cfDNA fragmentomic summaries supporting Figure 2 and Supplementary Figs. 2 and 3. Strict paired analyses used same-patient, same-timepoint PB/BM cfDNA pairs (n = 51), including high- and low-tumor-fraction strata (n = 34 and n = 17). Processing-context analyses stratified paired baseline samples as local/non-shipped (n = 11) or overnight-shipped (n = 40).

**Supplementary Table 3. Mutation threshold and tier framework summary.**

Candidate mutation-threshold frameworks and final tier definitions used for primary and exploratory cfDNA mutation analyses. The primary BM plasma-cell reference required VAF ≥ 0.10 and ALT ≥ 4. The primary Tier 1 cfDNA framework required VAF ≥ 0.0075, ALT ≥ 12, and DEPTH ≥ 500; the exploratory Tier 2 cfDNA framework required VAF ≥ 0.005, ALT ≥ 8, and DEPTH ≥ 250.

**Supplementary Table 4. BM-reference mutation threshold stability.**

Sensitivity of the BM plasma-cell mutation reference to local changes in BM-cell VAF, alternate-read, and depth thresholds. These analyses support the selected BM-reference threshold as a conservative clonal reference rather than a low-VAF marrow mutation catalogue.

**Supplementary Table 5. Post-calibration mutation-callset refinement impact.**

Impact of post-calibration refinement on baseline cfDNA mutation calls, including primary-call burden before and after refinement and pairwise BM-versus-cfDNA sharing. Demoted calls were reassigned to the exploratory tier rather than removed from the analysis set.

**Supplementary Table 6. Composition and quality of demoted/private mutation calls.**

Composition, caller support, VAF distribution, and oncogenicity characteristics of demoted and compartment-specific mutation calls. Denominators are mutation-call observations rather than patients unless otherwise specified.

**Supplementary Table 7. Subthreshold BM support for cfDNA-only Tier 1 mutations.**

Summary of matched BM plasma-cell support for cfDNA-only Tier 1 mutations that did not meet the BM-reference threshold. Denominators are cfDNA-only Tier 1 mutation observations by compartment and tumor-fraction stratum.

**Supplementary Table 8. Broader BM lookback evidence summary.**

Broader BM lookback evidence for BM-reference-discordant cfDNA mutation observations. Among 72 discordant cfDNA mutation observations, 15/72 (20.8%) showed broader but subthreshold BM evidence.

**Supplementary Table 9. BM lookback root-cause summary.**

Root-cause categorization of BM-reference-discordant cfDNA mutation observations, including BM truncation or subthreshold support, plausible biological heterogeneity, technical or caller-pattern concerns, and indeterminate evidence.

**Supplementary Table 10. CNA framework calibration and callability summary.**

Candidate CNA framework performance, selected CNA framework rationale, and lesion- or arm-level callability definitions. The selected primary CNA framework was cna_shared_alt0.5_cov0.4_tf0 (an altered-arm fraction of 0.5, a coverage fraction of 0.4, and did not impose a hard tumor-fraction exclusion). Denominators are endpoint- and lesion-specific callable event sets.

**Supplementary Table 11. Translocation calibration, FISH benchmarking, and event-set definitions.**

Candidate translocation-calling frameworks, selected-framework rationale, BM plasma-cell-referenced recovery metrics and orthogonal FISH benchmarking summaries. The selected primary framework, tx_shared_cf100_rescue400, used a BM plasma-cell clean-call cutoff of 40, a shared cfDNA clean-call cutoff of 100 and a conservative rescue score of 400. BM-reference translocation sensitivity used the strict BM-reference-positive denominator (n = 5); the broader event-class set (n = 6) is reported only where explicitly labeled. FISH benchmarking rows use patient-event counts and are distinct from the primary BM plasma-cell-referenced recovery population.

**Supplementary Table 12. BCR detection, calibration, and denominator definitions.**

BCR clonotype recovery and calibration summary with denominator categories kept distinct. The main-text BM-dominant callable detectability endpoint (31/34) and calibration-only target-callable threshold-output denominators (35/39 for BM plasma cfDNA and 37/41 for PB cfDNA) represent different analysis populations and should not be conflated.

**Supplementary Table 13. Targeted-panel design and capture content.**

Genomic coordinates, target annotations, and panel-component labels for the 257,803-bp custom hybrid-capture design used for mutation, translocation, and BCR analyses. The table includes recurrently altered myeloma genes, canonical immunoglobulin/MYC translocation hotspot regions, and IGH, IGK, and IGL V(D)J target regions. Coordinates are reported relative to GRCh38.

## Supplementary Notes

### Supplementary Methods

Calibration analyses were restricted to baseline samples with matched BM plasma-cell and cfDNA data for the relevant assay class. Denominators therefore varied by assay, compartment, callability, and availability of orthogonal FISH or BCR repertoire data. Each calibration endpoint therefore used the maximal evaluable subset for that assay.

For each assay class, cfDNA calls were compared with the BM plasma-cell reference only when both compartments were evaluable. Sensitivity represented the fraction of BM plasma-cell-positive events recovered in cfDNA. Specificity represented the fraction of BM plasma-cell-negative tests that remained negative in cfDNA. Balanced accuracy was the mean of sensitivity and specificity. CNA and translocation metrics were calculated across recurrent myeloma lesion or event tests. Mutation threshold metrics in Extended Data Fig. 4A-B used the evaluable comparison units defined in the source calibration tables, while variant-level event counts were used for event-architecture and cfDNA-only summaries.

#### Cohort- and sample-type-specific assay implementation

For TFRIM4 patients processed before 2023, CD138-enriched BM-cell DNA was processed through the Advanced Molecular Diagnostics Laboratory (AMDL) at Princess Margaret Hospital using the 37-gene panel only and sequenced at AMDL on an Illumina NextSeq 550. For these patients, matched cfDNA samples and longitudinal timepoints were subsequently profiled using separate targeted assays: the 37-gene panel for mutation analysis and the immunoglobulin/translocation panel for translocation and BCR analyses. From 2023 onward, newly processed TFRIM4 patients were profiled using the combined full-panel design, including gene, translocation-hotspot, and immunoglobulin V(D)J targets, and were sequenced on a NovaSeq 6000 where material was available. For older TFRIM4 patients with at least one sample already processed using separate panel components, subsequent samples were kept on the same assay configuration and sequencing strategy, where possible, to minimize within-patient technical heterogeneity. In IMMAGINE, BM-cell DNA was processed through AMDL using the 37-gene panel only and sequenced on a NextSeq 550; insufficient remaining material precluded immunoglobulin/translocation profiling of IMMAGINE BM-cell specimens. Consequently, IMMAGINE BM-cell data contributed to mutation-based marrow-reference analyses but not to BM-cell-referenced translocation or BCR analyses. IMMAGINE cfDNA and ALGONQUIN samples were profiled using the combined full-panel design where material permitted.

#### Fragmentomic compartment classification

Within each leave-one-patient-out training fold, numeric features were median-imputed, centered, and scaled using training-set parameters only. These parameters were then applied unchanged to the held-out samples. AUC confidence intervals were estimated using DeLong’s method, with bootstrap confidence intervals computed as a sensitivity analysis. For the primary permutation analysis, one-sided empirical P values were calculated as (*b* + 1)/(*B* + 1), where *b* was the number of permuted AUCs greater than or equal to the observed AUC and *B* = 999 valid permutations. Tumor-fraction subset analyses used the same leave-one-patient-out framework after stratifying samples by shallow-WGS tumor fraction, defining low tumor fraction as <5% and high tumor fraction as ≥5%. Feature importance was summarized from the absolute standardized coefficients of a logistic-regression model fit to the full analysis set after applying the same preprocessing scheme. Study-stratified performance was summarized by applying cohort labels to the leave-one-patient-out predictions and estimating cohort-specific ROC curves, AUCs, and within-patient differences in predicted BM plasma cfDNA probability between the two compartments.

For processing-context sensitivity analyses, matched baseline PB/BM plasma cfDNA pairs were classified according to BM sample processing context. For each selected fragmentomic feature, paired BM-minus-PB deltas were calculated within each patient’s matched baseline samples. Within each processing group, paired standardized effect sizes were summarized as paired Cohen’s dz, with patient-bootstrap 95% confidence intervals. Local-versus-shipped differences in patient-level paired deltas were assessed using Wilcoxon rank-sum tests with Benjamini-Hochberg adjustment across the tested features. These analyses were interpreted descriptively because processing context was partially confounded with cohort and collection site and because subgroup sample sizes were limited.

#### Mutation threshold calibration and BM-reference stability

Somatic mutation calibration used BM plasma cells as the clonal reference and evaluated cfDNA mutation thresholds separately from the BM-reference threshold. The retained BM plasma-cell reference required VAF >= 0.10 and at least 4 alternate reads. This conservative reference was an operational clonal anchor for benchmarking recovery of clonal marrow events; it was not intended to represent the full low-VAF marrow tail or all spatial disease biology.

Candidate cfDNA thresholds were screened across VAF, alternate-read, and depth cutoffs. The source threshold grid used VAF cutoffs of 0.001, 0.0025, 0.005, 0.0075, and 0.010; alternate-read cutoffs of 4, 6, 8, 10, and 12; and depth filters of 0, 250, 500, 1000, and 1500 (Extended Data Fig. 4A-B; Supplementary Table 3). A depth filter of DEPTH >= 0 indicated no additional minimum-depth filter beyond upstream mutation QC and evaluable-site requirements; it did not indicate that sites without meaningful sequencing coverage were considered evaluable. Candidate thresholds were evaluated by BM plasma-cell-referenced sensitivity, specificity, balanced accuracy, driver-mutation sensitivity, and the proportion of cfDNA-only calls. Driver mutations were defined as mutation events involving genes in a prespecified myeloma driver-gene framework used for the main mutation analyses (including *KRAS, NRAS, BRAF, TP53, DIS3, TENT5C, FAM46C, CYLD, TRAF3, TRAF2, CCND1, IRF4, PRDM1, RB1, ATM, ATR, KDM6A, FGFR3, MAX,* and *NFKB2*).

The cfDNA-only event ratio counted how many calls appeared only in one cfDNA compartment and not in the retained BM plasma-cell reference, defined as (BM plasma cfDNA-only + PB cfDNA-only events) divided by the union of BM plasma-cell, BM plasma cfDNA, and PB cfDNA events in that threshold setting. This metric was used to flag thresholds that gained calls mainly by adding cfDNA-only events, not as a formal false-discovery rate.

We chose the primary cfDNA threshold (VAF ≥ 0.0075; ALT ≥ 12; Depth ≥ 500) because it maximized balanced accuracy (0.830) against the bone marrow reference, while maintaining high mean specificity (0.986) and a lower cfDNA-only event-burden ratio than more permissive alternatives. The lower-support exploratory tier (VAF >= 0.005, ALT >= 8, DEPTH >= 250) had similar mean balanced accuracy (0.825) but a higher cfDNA-only event-burden ratio (0.674 versus 0.567 for Tier 1), so it was retained to preserve sensitivity-oriented observations for audit and sensitivity analyses but excluded from primary concordance, detectability, and clinical association claims (Extended Data Fig. 4A-E; Supplementary Table 3).

BM-reference stability was assessed by locally varying the BM plasma-cell VAF, alternate-read, and depth requirements while holding the selected cfDNA Tier 1 threshold fixed. The stability analysis examined the number of BM-reference events and drivers, plasma-compartment sensitivity, and event-level and driver-level Jaccard similarity relative to the retained BM reference. This supported the retained BM-reference definition as a stable operational anchor for cross-compartment comparisons, while recognizing that changes in reference thresholds naturally altered the number of evaluable mutations (Extended Data Fig. 4F; Supplementary Table 4).

#### cfDNA mutation tiering and post-calibration refinement

Mutation candidates were generated from the targeted-panel variant-calling ensemble used in the main analysis, including MuTect2, Strelka, VarDict, MuTect, and SomaticSniper, followed by annotation and rule-based tiering. BM plasma-cell calls meeting the BM-reference threshold (10% VAF) were assigned to the BM-reference callset; calls below this threshold were not treated as reference-positive events. For cfDNA, variants meeting the primary cfDNA threshold were assigned to Tier 1 unless they matched a post-calibration demotion rule. Variants below Tier 1 but meeting the lower-support cfDNA threshold were assigned to the exploratory Tier 2 callset.

After threshold selection, we applied a narrow refinement step to reduce recurrent low-confidence cfDNA-only patterns in the primary callset. Two non-oncogenic cfDNA patterns were moved from Tier 1 to Tier 2 when they also met Tier 2 support: non-oncogenic MuTect2-only indel-like calls and Strelka + VarDict missense calls lacking MuTect2 support. These calls were retained as exploratory observations rather than removed, preserving reproducibility and allowing sensitivity review while preventing recurrent low-confidence cfDNA-only patterns from driving primary concordance analyses (Extended Data Fig. 5A-B; Supplementary Tables 5 and 6).

Residual Tier 1 cfDNA-only mutations, defined as Tier 1 cfDNA calls absent from the BM plasma-cell reference, were then reviewed against the matched BM plasma-cell data. First, we asked whether the same variant was present in BM plasma cells below the 10% VAF reference cutoff. For the remaining discordant calls, we grouped the BM evidence by strength and interpretation: very-low-VAF support, low-VAF support, subclonal 5-10% support, no BM evidence but oncogenic plausibility, no BM evidence but non-oncogenic multi-caller support, single-caller support, or residual uncertainty. This review ensured that variants missing from the high-confidence marrow reference were not automatically dismissed as biologically absent. It also separated three possible explanations for cfDNA-only calls: exclusion by the BM-reference threshold, spatial or compartmental heterogeneity, and technical noise (Extended Data Fig. 5C-E; Supplementary Tables 7-9).

Baseline SNP trinucleotide-context and SBS-class summaries were used as callset-composition checks, not as reclassification rules. These summaries assessed whether Tier 1 or Tier 2 SNP callsets showed major differences in substitution patterns that could suggest systematic artefactual enrichment. They were not used to upgrade or downgrade individual mutation calls (Extended Data Fig. 5F-G).

#### CNA framework calibration

CNA calibration started from shallow whole-genome sequencing segmentation and ichorCNA-derived tumor-fraction estimates, then translated segment-level results into recurrent myeloma arm-level lesions. Candidate CNA frameworks varied the altered-arm fraction threshold, arm coverage requirement, and tumor-fraction masking rules. Frameworks were compared using BM plasma-cell-referenced sensitivity, specificity, balanced accuracy, orthogonal FISH specificity where available, cfDNA-only event inflation, and retained callability. Framework-level CNA metrics were averaged across the recurrent myeloma arm-level lesion tests evaluated in the calibration grid.

The selected CNA framework (cna_shared_alt0.5_cov0.4_tf0) required an altered-arm fraction of 0.5, a coverage fraction of 0.4, and did not impose a hard tumor-fraction exclusion for categorical CNA calls. This framework was selected for its balance of performance and reliability. Although other candidates showed marginally higher accuracy, they significantly limited the number of samples that could be analyzed (reducing callability from 0.655 to 0.319). A hard tumor-fraction mask was therefore not selected because it reduced callability without producing a sufficient gain in balanced accuracy or specificity in the calibration grid. The selected rule was retained as a single shared primary framework across BM plasma cfDNA and PB cfDNA rather than a compartment-specific or tiered CNA framework (Extended Data Fig. 6A-B; Supplementary Table 10).

#### Translocation framework calibration and FISH benchmarking

Canonical myeloma translocations were called from targeted-panel data with IgCaller, filtered for caller support, blacklist status, immunoglobulin-side plausibility, and proximity to canonical myeloma partner loci. Candidate frameworks varied clean-call score cutoffs, shared versus compartment-specific cfDNA thresholds, and rescue settings for highly supported near-canonical events. Framework selection prioritized BM plasma-cell-referenced balanced accuracy, FISH specificity, and control of cfDNA-only inflation. The selected framework (tx_shared_cf100_rescue400) used a shared cfDNA clean-call cutoff of 100, retained a BM plasma-cell cutoff of 40, and allowed a conservative rescue score of 400. The no-rescue cf100 framework had the highest BM-reference balanced accuracy, but it had lower FISH-positive recovery; the selected conservative rescue rule recovered more FISH-supported events while keeping specificity high and cfDNA-only burden lower than more permissive rescue or lower-score cfDNA thresholds. A single conservative primary translocation callset was used rather than a tiered design because the number of evaluable translocation events was small and permissive thresholds disproportionately increased cfDNA-only calls (Extended Data Fig. 6C-D; Supplementary Table 11).

FISH benchmarking was treated as an orthogonal clinical-assay audit rather than a replacement truth set, because FISH and targeted-panel calls do not always interrogate identical molecular breakpoints or copy-number boundaries. For each evaluable CNA or translocation event, sensitivity was calculated among FISH-positive cases and specificity among FISH-negative cases, separately for BM plasma cells, BM plasma cfDNA, and PB cfDNA. Sparse FISH denominators were explicitly marked because several translocation and lesion-specific comparisons had fewer than three positive events (Extended Data Fig. 6E; Supplementary Table 11).

#### BCR dominant-clonotype calibration

BCR calibration used productive immunoglobulin clonotypes defined by locus, CDR3 amino-acid sequence, and top V/J assignments. For BM plasma-cell-referenced dominant-clonotype recovery, a BM plasma-cell clonotype qualified as the reference only if the top productive clonotype at that locus had at least 5 reads, at least 10% of productive reads, no top-read tie, and at least 1.5x dominance over the runner-up when present. Among 42 baseline patients with BM cell BCR data available, 36 had a qualifying BM-dominant clonotype detected.

We then attempted to detect the BM-dominant BCR in matched cfDNA. In the cfDNA target compartment, the matched locus was considered callable when it had at least 10 productive reads, and the BM-matched clonotype was considered recovered when at least 5 target reads matched the BM-dominant clonotype. This target rule was selected because it removed most 1-4 read edge cases while preserving high same-patient recovery (35/39 total callable BM plasma cfDNA baseline and 37/41 PB cfDNA target-callable plasma cfDNA cases) and low same-locus other-patient decoy match rates (0.10% for BM plasma cfDNA and 0.07% for PB cfDNA). Of the 36 patients with a BM-dominant BCR reference, 34 had callable BM plasma cfDNA, and 34 had callable PB cfDNA for this endpoint. Matched clone fraction was retained for documentation but was not used as the primary gate because fractions are unstable at very low read depth (Extended Data Fig. 6F-H; Supplementary Table 12).

### Supporting Calibration Results

#### Fragmentomic compartment classification was evident across tumor-fraction strata and study contexts

To complement the univariate fragment-size and end-motif analyses in the main text, we evaluated whether these features formed a coherent multifeature compartment signature. A leave-one-patient-out logistic-regression classifier trained on fragment-size and 5′ end-motif summary features distinguished baseline BM plasma cfDNA from matched PB cfDNA (LOOCV AUC, 0.910; 95% CI, 0.842–0.978; within-patient label-permutation P = 0.001; Extended Data Fig. 3C). Performance was retained in low- and high-tumor-fraction subsets (AUC, 0.856 and 0.872, respectively), supporting compartment separability that was not driven only by high tumor-fraction samples.

Feature-importance analysis showed that classifier separation was driven primarily by long-fragment fraction, insert-size entropy, median insert size, and 5′ end-motif features, whereas short-fragment fraction, mono-/di-nucleosome ratio, and the myeloma-region 5-Mb fragmentation-ratio summary contributed less strongly (Extended Data Fig. 3D). Study-stratified analyses showed that BM plasma cfDNA generally received higher predicted BM-compartment probabilities than matched PB cfDNA across contributing cohorts, with the weakest separation in IMMAGINE (Extended Data Fig. 3E,F). These findings support a multifeature fragmentomic signature distinguishing BM plasma cfDNA from PB cfDNA, while also indicating that the magnitude of separation varies by study context.

#### Mutation calibration supports a conservative primary tier and a separate exploratory tier

The shared Tier 1 threshold was selected because it balanced BM plasma-cell-referenced accuracy while limiting cfDNA-only calls. Under the selected Tier 1 rule, balanced accuracy was 0.820 for BM plasma cfDNA and 0.840 for PB cfDNA in BM plasma-cell-referenced comparisons (Extended Data Fig. 4A-C; Supplementary Table 3). Differences in apparent BM plasma cfDNA and PB cfDNA balanced accuracy were not interpreted as evidence that one cfDNA compartment performed better, because assay availability, cfDNA quality, tumor fraction, and event callability differed across evaluable subsets. A compartment-specific Tier 1 framework provided only marginal benefit and was not adopted because it added interpretive complexity without materially changing the conclusions. Tier 2 preserved lower-support observations for sensitivity review, but primary concordance, detectability, and clinical claims were anchored to Tier 1 (Extended Data Fig. 4A-E; Supplementary Table 3).

Mutation calls were categorized by detection pattern across compartments, including BM plasma-cell-reference only, BM plasma-cell plus BM plasma cfDNA, BM plasma-cell plus PB cfDNA, all-three-compartment, BM plasma cfDNA plus PB cfDNA without BM-reference support, BM plasma cfDNA-only, and PB cfDNA-only categories, as applicable to the plotted threshold tiers (Extended Data Fig. 4D). The retained BM plasma-cell reference contained 28 mutation events, including 25 driver events across 23 patients. Local BM-reference threshold variation around VAF ≥ 0.10 and ALT ≥ 4 left the retained reference unchanged for nearby alternate-read and depth changes, while raising the BM VAF threshold to 0.125 reduced the reference to 24 events and 23 drivers. These results support the retained BM-reference threshold as a locally stable operational clonal anchor, not as an exhaustive representation of low-VAF marrow biology (Extended Data Fig. 4F; Supplementary Table 4).

#### cfDNA mutation refinement reduced cfDNA-only inflation but did not erase biological ambiguity

Post-calibration refinement moved 9/64 BM plasma cfDNA calls (14.1%) and 25/76 PB cfDNA calls (32.9%) from the previous primary cfDNA mutation callset to the exploratory tier (Extended Data Fig. 5A; Supplementary Table 5). In PB cfDNA, demoted calls consisted of 23 Strelka + VarDict non-oncogenic missense calls lacking MuTect2 support and 2 MuTect2-only non-oncogenic frame-shift deletions. In BM plasma cfDNA, demotions primarily consisted of MuTect2-only non-oncogenic indel-like calls, including 6 frame-shift deletions, 2 in-frame deletions, and 1 in-frame insertion (Extended Data Fig. 5B; Supplementary Table 6). The higher demotion fraction in PB cfDNA reflected enrichment of the recurrent non-oncogenic missense pattern supported by Strelka and VarDict but not MuTect2, rather than a broad loss of PB cfDNA signal.

BM lookback analyses showed that cfDNA-only mutations were not always completely absent from marrow. Among matched samples, 9/12 PB cfDNA-only Tier 1 mutations and 5/23 BM plasma cfDNA-only Tier 1 mutations had exact subthreshold BM plasma-cell support below the 10% VAF reference cutoff (Extended Data Fig. 5C-D; Supplementary Table 7). In the broader lookback, 15/72 discordant cfDNA mutation observations (20.8%) had detectable but subthreshold BM evidence, including 2 very-low-VAF, 7 low-VAF, and 6 subclonal 5-10% BM observations (Extended Data Fig. 5D-E; Supplementary Tables 8 and 9). The remaining discordant calls included oncogenic cfDNA-only calls that could reflect biological heterogeneity, non-oncogenic calls supported by multiple callers, and single-caller calls (Extended Data Fig. 5E; Supplementary Table 9).

Trinucleotide-context and SBS-class summaries were used as additional mutation-composition checks and were not used to reclassify individual variants (Extended Data Fig. 5F-G). At the SBS6 level, Tier 1 spectra did not differ significantly between compartments (Fisher exact P = 0.703 for BM plasma cells versus BM plasma cfDNA, P = 0.865 for BM plasma cells versus PB cfDNA, and P = 0.447 for BM plasma cfDNA versus PB cfDNA). In the exploratory Tier 2 comparison, PB cfDNA showed enrichment for T>C substitutions relative to BM plasma cfDNA (overall Fisher exact P = 0.013; T>C BH-adjusted P = 0.0027) (Extended Data Fig. 5F-G). These findings suggest that cfDNA-only Tier 1 mutations may arise from spatial heterogeneity or technical noise. Consequently, they should not be automatically dismissed as false positives, nor should they be used to claim that cfDNA is universally superior to marrow profiling.

#### CNA calibration supported one shared framework for PB and BM plasma cfDNA

The selected CNA framework (cna_shared_alt0.5_cov0.4_tf0) used one shared rule for BM plasma cfDNA and PB cfDNA, with an altered-arm fraction of 0.5, a coverage fraction of 0.4, and no hard tumor-fraction mask. Across the calibration grid, this framework had a mean balanced accuracy of 0.803 using BM plasma cells as the reference, with mean sensitivity of 0.646, mean specificity of 0.959, and an evaluable-test fraction of 0.655, averaged across recurrent myeloma arm-level lesion tests (Extended Data Fig. 6A-B; Supplementary Table 10). Nearby candidate frameworks performed similarly. CNA calibration was therefore interpreted mainly as evidence that the selected shared rule was stable and retained sufficient evaluable tests, rather than evidence that a more complex tiered framework was needed.

Under the selected framework, PB cfDNA recovered 24/31 BM-reference-positive CNA events across 110 callable event tests, yielding 77.4% sensitivity and 98.7% specificity. BM plasma cfDNA recovered 18/25 BM-reference-positive CNA events across 90 callable event tests, yielding 72.0% sensitivity and 92.3% specificity (Extended Data Fig. 6A-B; Supplementary Table 10). Orthogonal FISH benchmarking supported high specificity for many lesion-compartment combinations, but positive denominators were sparse and performance varied by lesion, especially for individual high-risk lesions. Cells with sparse denominators in Extended Data Fig. 6E should therefore be interpreted cautiously (Extended Data Fig. 6E; Supplementary Table 11).

#### Translocation calibration was limited by small event numbers

Translocation calibration was informative but limited by the small number of BM-reference-positive translocation events. The selected translocation framework (tx_shared_cf100_rescue400) had framework-level mean balanced accuracy of 0.893 using BM plasma cells as the reference, with mean sensitivity of 0.625, mean specificity of 0.980, and FISH-any mean balanced accuracy of 0.915 across the calibration summaries (Extended Data Fig. 6C-D; Supplementary Table 11). The retained framework favored specificity over more permissive rescue rules, which increased cfDNA-only calls.

Final BM-reference recovery estimates should be considered descriptive because only 5 BM-reference-positive translocation events were available, with 6 events in the broader event class. In this small BM-reference-positive denominator, PB cfDNA recovered 5/5 translocations, and BM plasma cfDNA recovered 2/5. These estimates should not be interpreted as evidence of superior PB translocation sensitivity. Specificity was high in both compartments: 99.5% for PB cfDNA and 98.6% for BM plasma cfDNA across 222 callable event tests (Extended Data Fig. 6C-E; Supplementary Table 11). FISH benchmarking reinforced this caution because some event-level comparisons had no positive cases or fewer than three positive cases (Extended Data Fig. 6E; Supplementary Table 11).

#### BCR clonotype recovery was high but governed by target-locus callability

Extended Data Fig. 6F summarizes target-locus callability, Extended Data Fig. 6G summarizes read support for the BM-matched clonotype, and Extended Data Fig. 6H shows recovery stability across target-read thresholds. Locus-level calibration supported the chosen target-support thresholds. For BM plasma cfDNA, target-locus callability and recovery were 26/34 and 17/34 for IGH, 27/30 and 21/30 for IGK, and 18/23 and 15/23 for IGL. For PB cfDNA, the corresponding values were 27/35 and 24/35 for IGH, 28/31 and 24/31 for IGK, and 18/26 and 15/26 for IGL (Extended Data Fig. 6F-H; Supplementary Table 12). These results support BCR as a high-recovery clonotype readout while emphasizing that BCR sharing, BM-dominant clonotype recovery, and lesion-specific genomic detection are biologically and analytically different endpoints.

