## Extended Data Figure 1 for "Matched marrow and blood profiling reveals compartment- and feature-specific liquid-biopsy signals in multiple myeloma"

### Extended Data Fig. 1. Cohort-specific sample flow, assay availability, cfDNA yield, targeted-panel design, and BM plasma cfDNA processing context

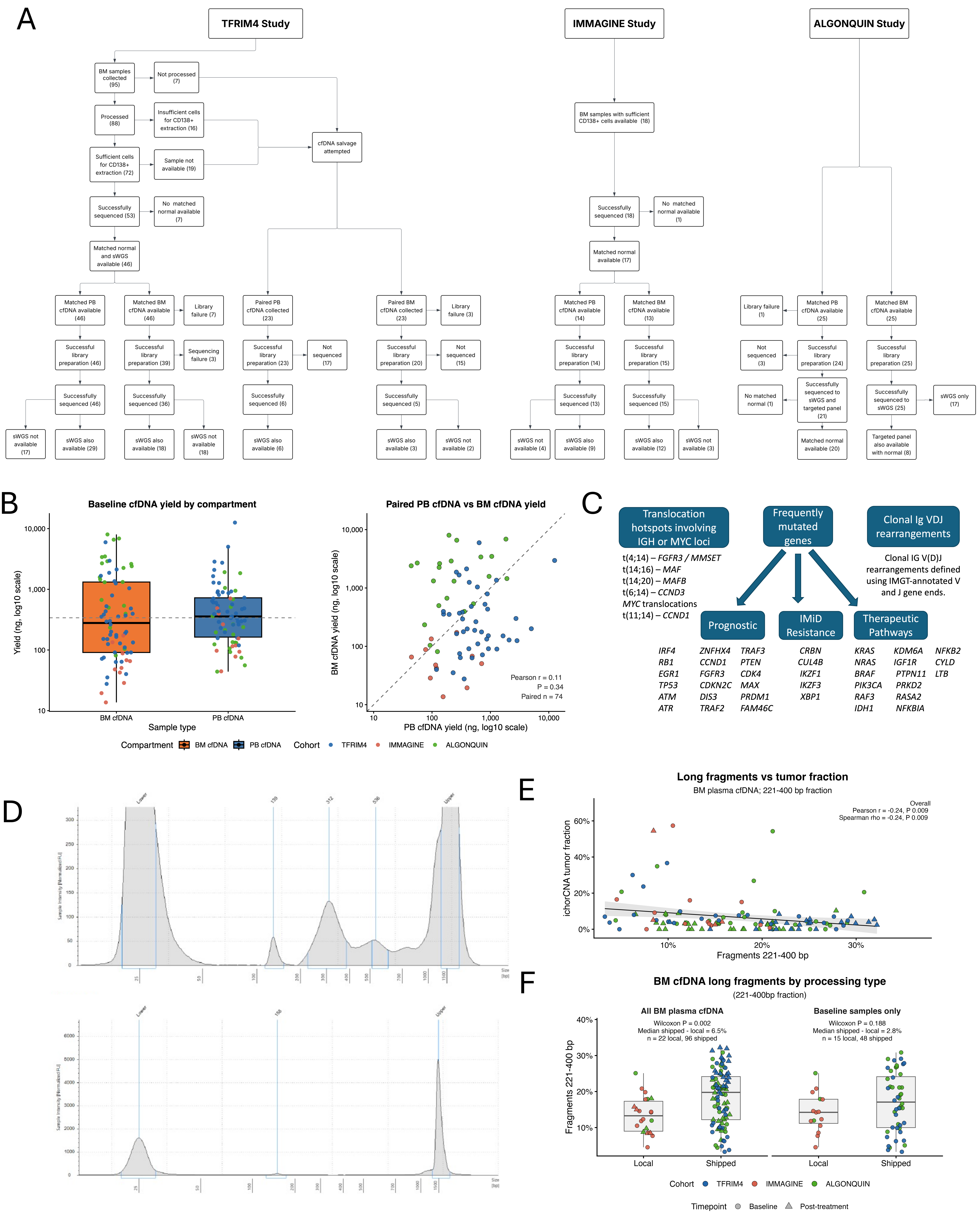
