## Extended Data Figure 2 for "Matched marrow and blood profiling reveals compartment- and feature-specific liquid-biopsy signals in multiple myeloma"

### Extended Data Fig. 2: Paired fragment-size, nucleosomal, and regional-fragmentation features in BM plasma cfDNA and PB cfDNA

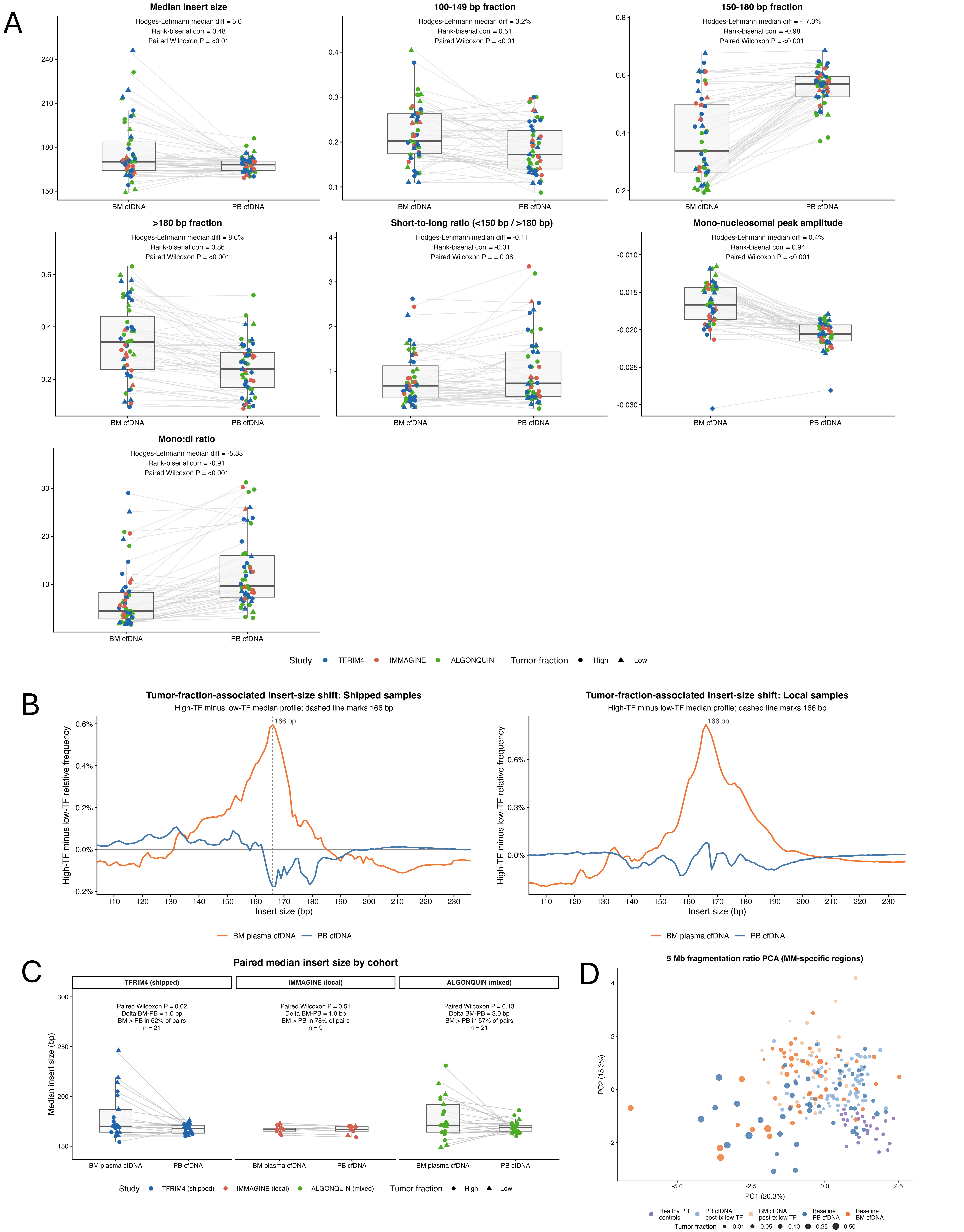
