## Extended Data Figure 3 for "Matched marrow and blood profiling reveals compartment- and feature-specific liquid-biopsy signals in multiple myeloma"

### Extended Data Fig. 3: End-motif and fragmentomic features support compartment- and processing-aware interpretation of BM plasma cfDNA and PB cfDNA

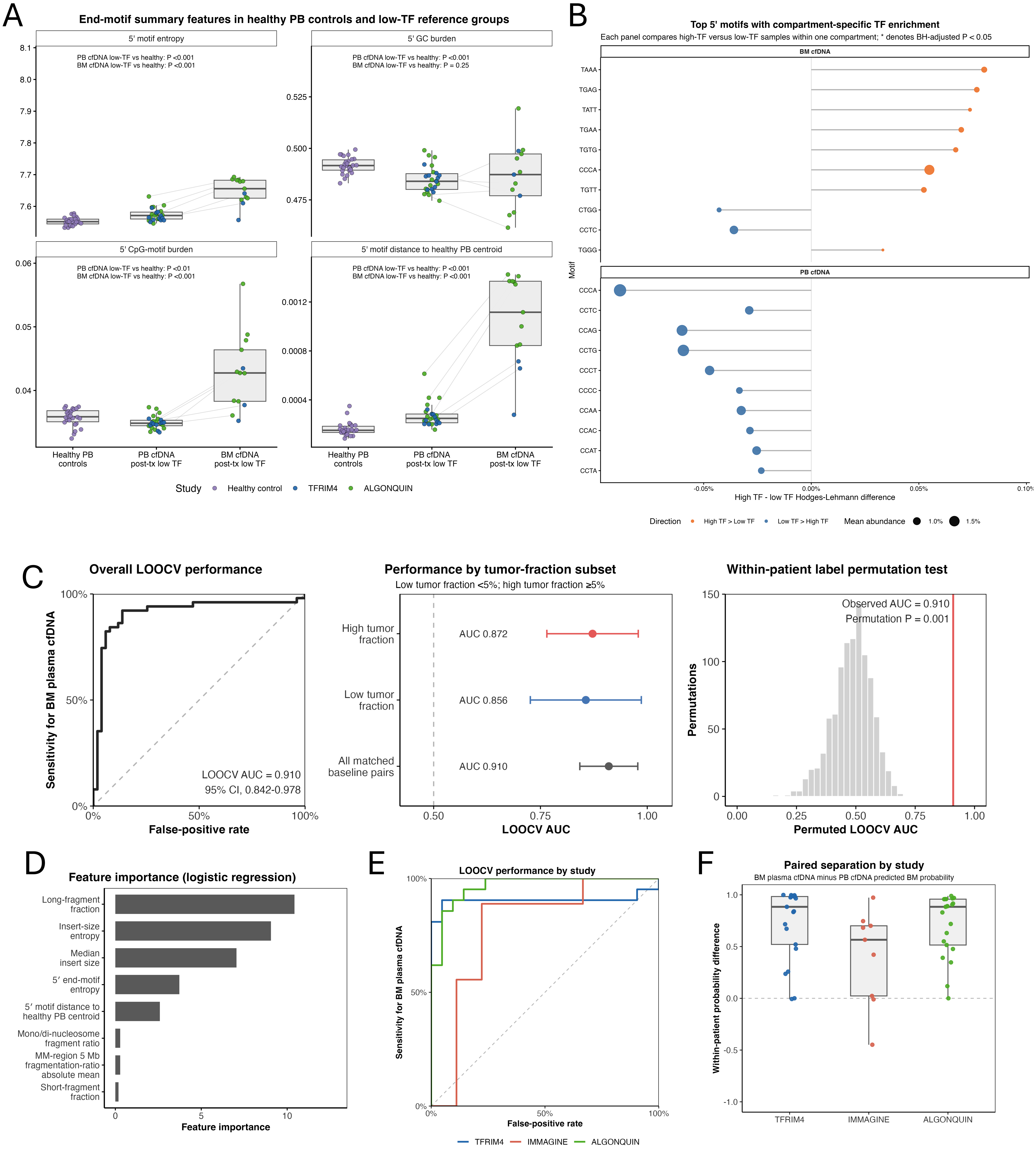
