## Extended Data Figure 4 for "Matched marrow and blood profiling reveals compartment- and feature-specific liquid-biopsy signals in multiple myeloma"

### Extended Data Fig. 4: Calibration and stability of BM plasma-cell-referenced cfDNA mutation thresholds

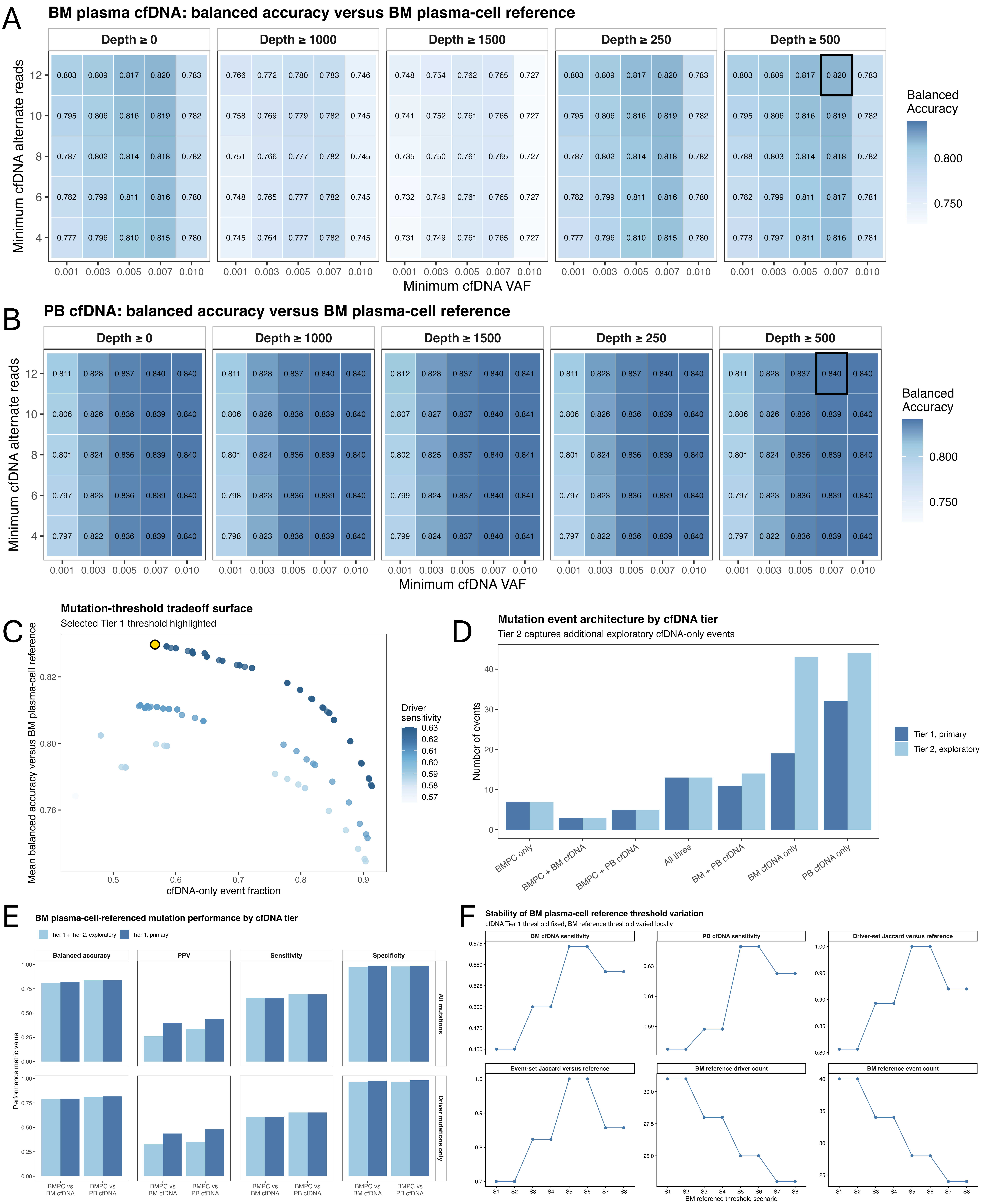
