## Supplementary figures and images for "Matched marrow and blood profiling reveals compartment- and feature-specific liquid-biopsy signals in multiple myeloma"

### Extended Data Figure 5

#### Extended Data Fig. 5: Post-calibration refinement and adjudication of cfDNA mutation calls

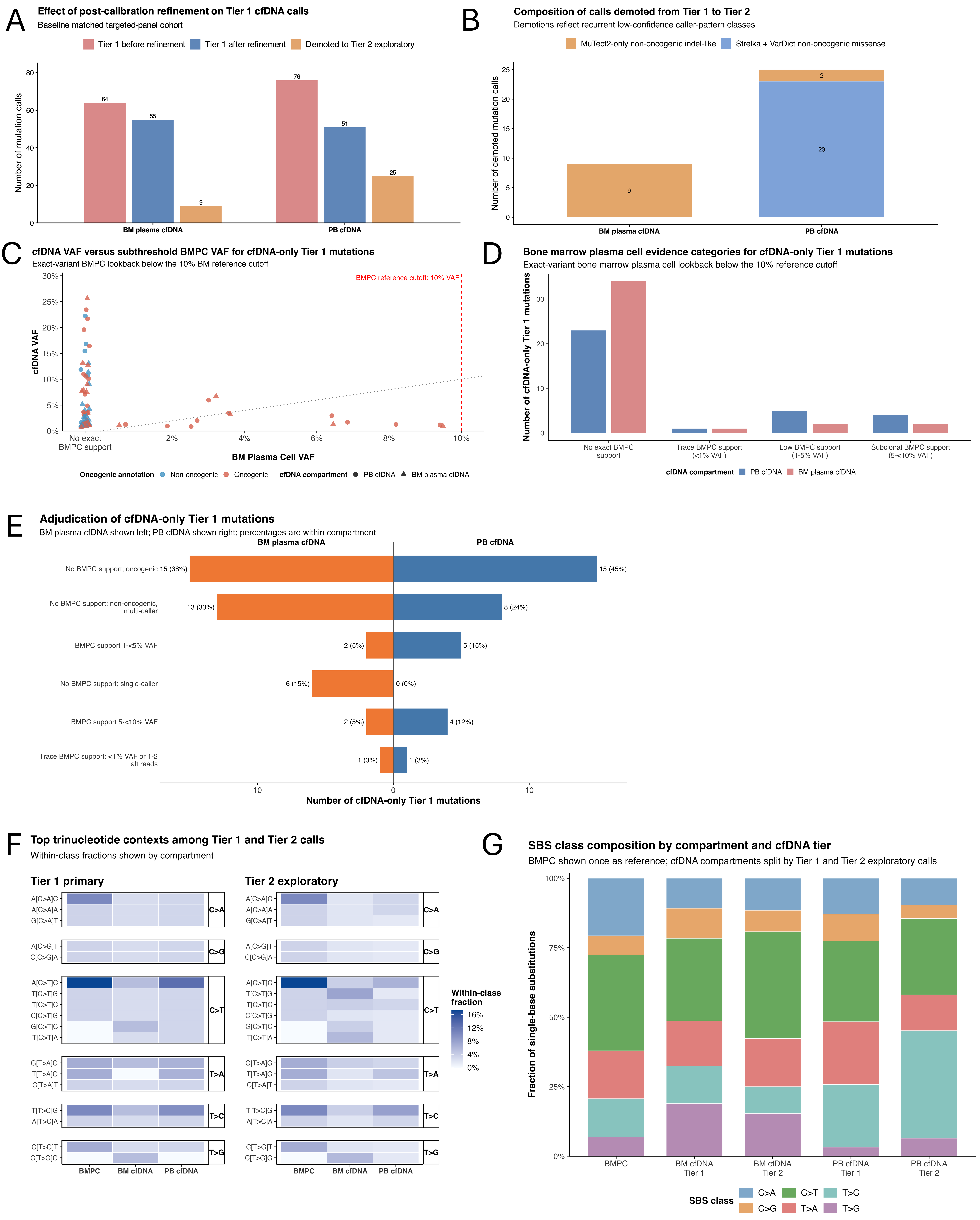
