## Extended Data Figure 6 for "Matched marrow and blood profiling reveals compartment- and feature-specific liquid-biopsy signals in multiple myeloma"

### Extended Data Fig. 6: Calibration and benchmarking of CNA, translocation, and BCR recovery in BM cfDNA and PB cfDNA

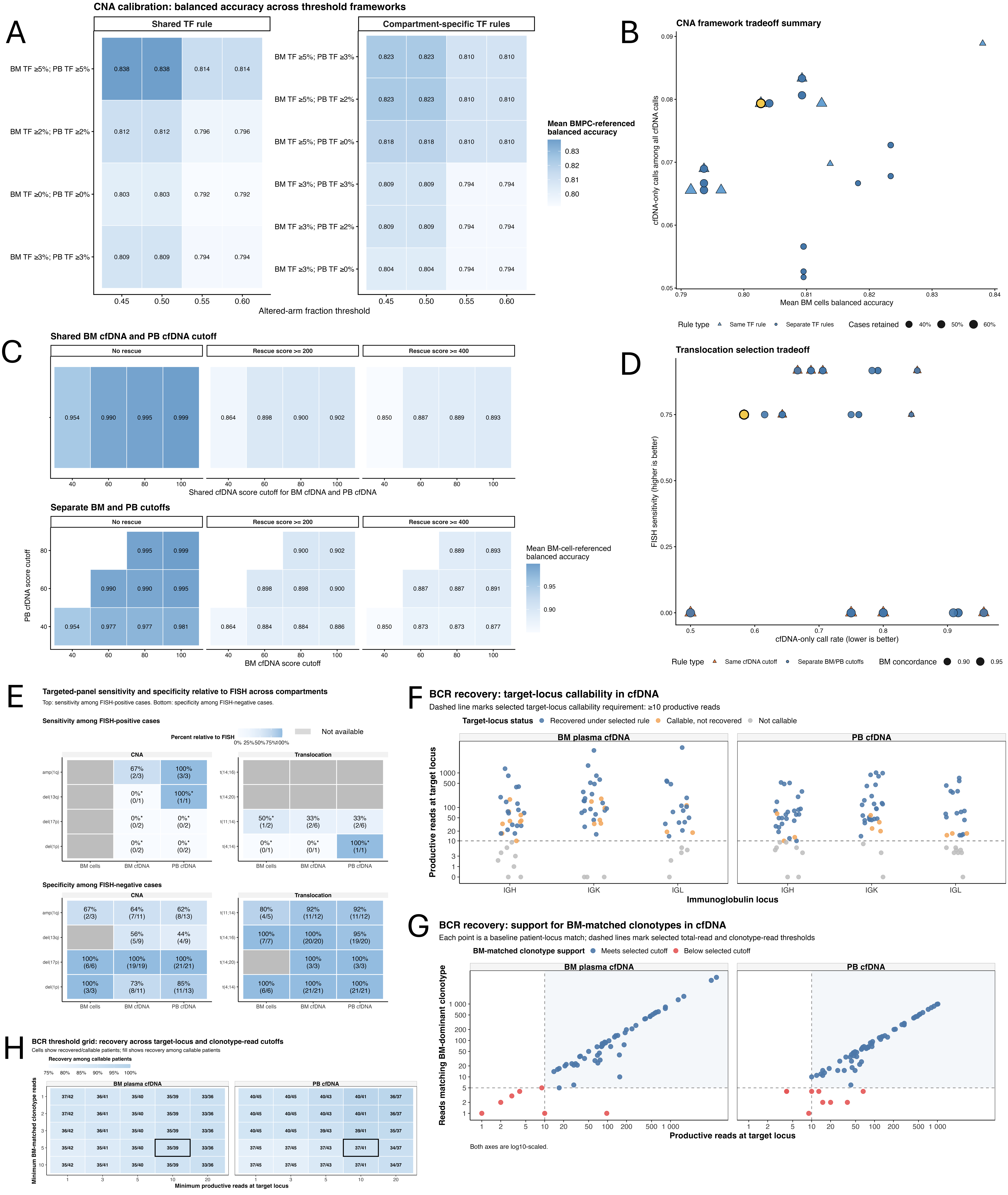
